# A mechanistic model for genetic regulation of postmenopausal bone loss

**DOI:** 10.64898/2026.06.04.26354968

**Authors:** Ilia Rattsev, Feilim Mac Gabhann, Daniel Hertz, Casey Overby Taylor

## Abstract

Bone remodeling is a tightly regulated physiological process that maintains bone health through coordinated action of bone-resorbing osteoclasts and bone-forming osteoblasts. Disruption of this balance, such as the one induced by estrogen decline after menopause, results in bone loss and osteoporosis. Genetic factors play an important role in determining bone mineral density (BMD) loss over time. However, translating genetic associations into individualized risk prediction remains challenging due to small effect size of individuals variants and non-linear interactions within the bone remodeling unit. Here, we present a bone cell population dynamics model that includes major regulatory pathways, such as the RANK/RANKL/OPG axis, Wnt signaling, and hormonal regulation by estrogen, parathyroid hormone, and TGF-*β*. We calibrate the model on clinical data from healthy postmenopausal women, and women with reduced BMD undergoing anti-osteoporotic therapy. The calibrated model captures healthy BMD decline in postmenopausal women and therapeutic response to anti-osteoporotic medications. We mechanistically incorporate the effect of 22 variants across 8 genes involved in bone remodeling and simulate BMD trajectories in 1,000 virtual subjects differing by ancestry and genetic makeup. The median predicted 5-year BMD loss was 3.57% (95% prediction interval: 1.31–5.24), consistent with the values reported in the literature. The virtual individuals with African ancestry were predicted to experience the highest average 5-year BMD loss. The strongest genetic risk factors for bone loss were predicted to be *CYP19A1* rs727479 and *OPG* rs3102735, while *LRP5* rs11228240 emerged as a protective factor that could partially counteract the detrimental effects of other variants. Several epistatic effects were observed in the genetic interaction analysis. Mechanistically, our model suggested that estrogen exerts its effect on bone remodeling primarily by modulating osteoclast apoptosis. Overall, this framework demonstrates a proof-of-concept for integration of genetic risk factors into mechanistic models of disease and can be extended to other conditions with polygenic inheritance.

## Introduction

Bone remodeling is a tightly regulated physiological process characterized by a continuous resorption of old or damaged bone by osteoclasts, followed by the deposition of new bone tissue by osteoblasts [1]. The balance between osteoclast and osteoblast activities is essential to maintain bone homeostasis [2]. Dysregulation of this process can result in bone metabolic disorders: excessive bone formation may lead to increased bone mineral density (BMD) and osteopetrosis, while excessive resorption may result in bone loss and osteoporosis [2]. Several molecular regulators and signaling pathways are crucial for maintaining bone homeostasis. Osteoclast differentiation and maturation is controlled by the receptor activator of nuclear factor kappa-B (RANK), its ligand (RANKL), and osteoprotegerin (OPG) [2, 3]. Osteoblast differentiation and activation is regulated by Wnt signaling and its modulators sclerostin and Dickkopf-1 (DKK-1) [2, 4, 5].

Osteoporosis is a common bone metabolic disorder, characterized by low BMD and an increased risk of fractures [6, 7]. It is more prevalent in older age and disproportionally affects women, with an estimated 200 million women affected worldwide [8]. It is well-established that increased bone remodeling in postmenopausal women is a direct consequence of estrogen decline [7, 9]. Estrogen plays a key role in maintaining bone homeostasis by regulating osteoclast apoptosis and modulating sclerostin, RANKL, and OPG levels [10–15]. After the menopause, estrogen levels drop dramatically, resulting in increased bone resorption, often leading to postmenopausal osteoporosis [7].

Several mathematical models of bone remodeling describe the complex interplay between osteoclasts, osteoblasts, and the regulatory factors within the bone multicellular unit (BMU). The foundational works by Lemaire et al. (2004) and Pivonka et al. (2008) established frameworks for describing bone cell population dynamics, which were subsequently extended to incorporate additional biological complexity [16–19]. However, relatively few previously published models explicitly incorporated estrogen signaling, limiting a comprehensive assessment of postmenopausal bone dynamics [20–25]. In addition, existing models do not enable personalized risk assessment, as they do not incorporate individual-level variables.

Osteoporosis is a highly heritable disorder, with heritability estimates ranging from 50 to 85% [26, 27]. Several candidate gene and genome-wide association studies (GWAS) have identified single nucleotide polymorphisms (SNPs) associated with reduced BMD, osteoporosis diagnosis, and fracture risk [27–31]. However, BMD is a highly polygenic trait, and individual SNPs have small effect sizes. In addition, the complex interplay between cellular and molecular regulators of bone remodeling creates non-linear interactions between SNPs, making it challenging to translate genetic associations into clinically meaningful predictions.

In this study, we introduce a novel mechanistic model of bone remodeling that integrates estrogen signaling and genetic polymorphisms associated with BMD or estrogen levels in postmenopausal women. Building on the models by Farhat et al. (2017) and Jörg et al. (2022), we expand on previous work by explicitly incorporating estrogen effect osteoclast apoptosis and expression of RANKL, OPG and sclerostin [22, 32]. We simulate healthy bone dynamics in postmenopausal women, model the response to therapeutic interventions, and evaluate how different genetic profiles may affect the risk of bone loss in this population.

## Materials and methods

### Model structure

Our model structure is based on the bone cell population dynamics model originally proposed by Lemaire et al.(2004) and further extended by Pivonka et al. (2008), Farhat et al. (2017) and Jörg et al. (2022) [16, 17, 22, 32]. These models assume that change in BMD over time is proportional to the number of active osteoclasts and osteoblasts in relation to one another. One osteoclast unit is assumed to resorb the bone with a resorption rate constant *k res*, and one osteoblast unit to deposit new bone with a formation rate constant *k form*. In homeostatic state, these two processes are in equilibrium, and there is no net change in BMD over time. The numbers of active osteoclasts and osteoblasts are ultimately determined by their rates of differentiation from the progenitor cells and apoptosis rates. These processes are in turn regulated by a complex molecular network within the bone multicellular unit (BMU). In particular, RANK/RANKL/OPG axis regulates osteoclast differentiation, while Wnt signaling modulates osteoblast activity. To better understand the bone dynamics in postmenopausal women, in addition to the processes included in the previous models, we explicitly model regulation of RANKL, Wnt signaling, and osteoclast apoptosis by estrogen. The schematic representation of the model structure is presented in Figure 1.

**Fig 1.**
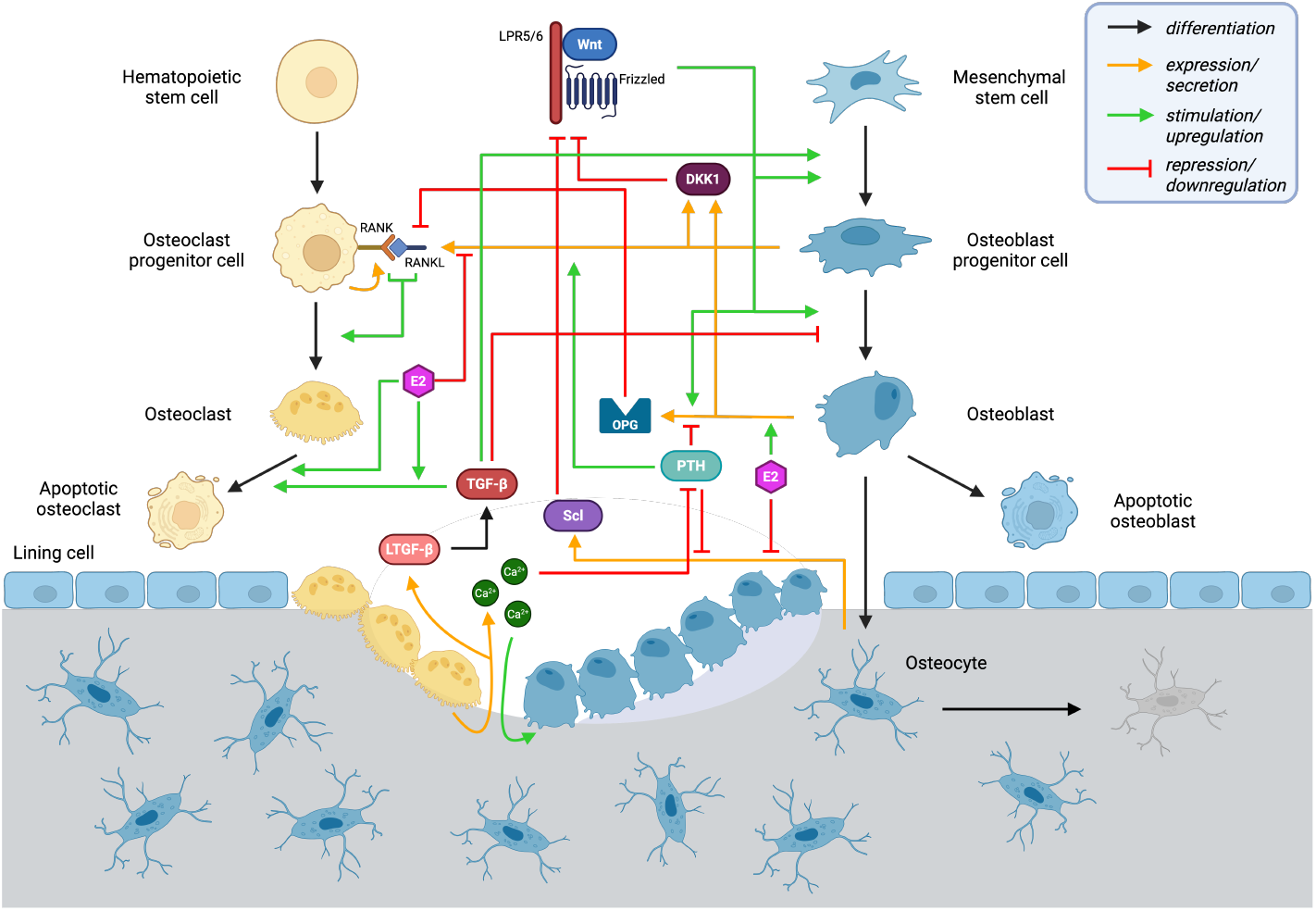
Bone remodeling model diagram. The model describes the dynamics of the bone multicellular unit (BMU) during bone remodeling. Bone is resorbed by osteoclasts and formed by osteoblasts. Black arrows indicate the transitions between cell states, such as differentiation and apoptosis. Orange arrows represent secretion of molecular species. A green arrow originating from a species indicates that the species stimulates the process to which it is pointing. A red inhibitory arrow originating from a species represents downregulation of the process to which it is pointing. *Ca*^2+^ *– calcium ions; DKK1 – Dickkopf-1; E2 – stradiol; LRP5 – low-density lipoprotein receptor-related protein 5; OPG – osteoprotegerin; PTH – parathyroid hormone; RANK – receptor activator of nuclear factor κB; RANKL – RANK ligand; Scl – sclerostin; TGF-β – transforming growth factor β; LTGF-β – latent transforming growth factor β*

### Mathematical formulation

The cellular and molecular dynamics were modeled using a system of ordinary differential equations (ODEs). Regulatory mechanisms were assumed to be mediated by receptor-ligand interactions controlling protein expression and cellular fate decisions, and were described using Hill-type functions of the form:

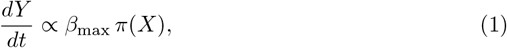

where *β*_*max*_ is the maximal reaction rate and *π* is the activator or repressor function that describes the effect of the regulatory factor *X* on concentration of *Y* .

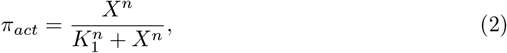

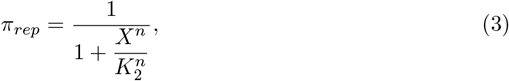

where *π*_*act*_ and *π*_*rep*_ are Hill functions for activator and repressor, respectively; *K*_1_ and *K*_2_ are activation and repression coefficients; and *n* is the Hill coefficient that indicates degree of cooperativity in enzyme-substrate binding. Unless specified otherwise, we assume independent binding and thus, *n* = 1. When multiple species *X* and *Z* independently regulate process *β*, we adopt the *OR* logic to calculate the combined regulatory effect and subtract the interaction term:

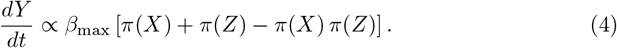

Conversely, when a synergistic effect exists between the two regulators, the interaction term is not subtracted:

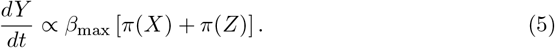

The system of ODEs was first equilibrated to steady state and then solved numerically in MATLAB (R2024b) using the stiff ODE solver *ode15s*.

#### Osteoclast progenitor cells (*OC*_*p*_)

Osteoclasts originate from hematopoietic stem cells (HSCs) that differentiate into osteoclast precursors, which fuse together to form mature osteoclasts [33]. We assume that the number of available HSCs is much larger than the number of osteoclast progenitors (*N*_*HSC*_ → ∞), such that osteoclast progenitors are generated at a constant HSC differentiation rate, 𝒟_*HSC*_. Osteoclast progenitors express receptor activator of nuclear factor *κ*B (RANK). Binding of RANK to its ligand, RANKL, stimulates the differentiation and maturation of osteoclast precursors. The populations of osteoclast progenitors can then be described with the following equation:

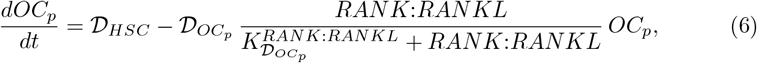

where *OC*_*p*_ is the osteoclast progenitor density within the BMU; 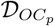 is the maximal differentiation rate of osteoclast progenitors; *RANK*:*RANKL* represents the concentration of the RANK-RANKL complex; and 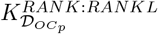 is the corresponding activation coefficient for RANK-mediated differentiation.

#### Active osteoclasts (*OC*_*a*_)

Active osteoclasts are formed by differentiation of osteoclast progenitors, which is stimulated by RANK-RANKL binding. Active osteoclasts undergo apoptosis at a maximal rate 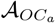. Estrogen and TGF-*β* signaling stimulate osteoclast apoptosis [22, 34]. Estrogen has been suggested to induce osteoclast apoptosis both directly via estrogen receptor-*α* (ER*α*) signaling, and via interaction with TGF-*β* [10, 22, 35]. Therefore, the population of active osteoclasts (*OC*_*a*_) can be described as

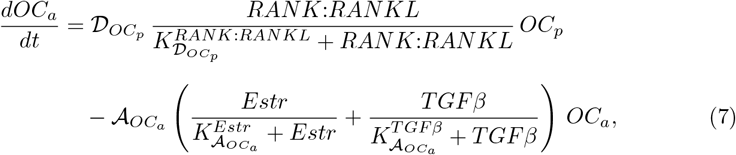

where *Estr* is the estradiol concentration; *TGFβ* is the TGF-*β* concentration; and 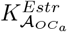 and 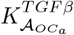 are the corresponding activation coefficients for estradiol- and TGF-*β*-mediated osteoclast apoptosis. Since estrogen’s effect on osteoclast apoptosis is partially mediated through TGF-*β* signaling, we do not subtract the interaction term between the two effects.

#### Osteoblast progenitor cells (*OB*_*p*_)

Osteoblast progenitor cells originate from a pool of mesenchymal stem cells (MSCs). We assume that the number of available MSCs is much larger than the number of osteoblast precursors (*N*_*MSC*_→ ∞), such that the rate of osteoblast precursor formation, 𝒟_*MSC*_, is independent of MSC abundance. Differentiation of MSCs into osteoblast progenitor cells is stimulated by both TGF-*β* and canonical Wnt signaling [32]. Canonical Wnt signaling further promotes the differentiation of osteoblast progenitors into active osteoblasts, whereas TGF-*β* inhibits this process [32]. The following equation describes the dynamics of osteoblast progenitors (*OB*_*p*_):

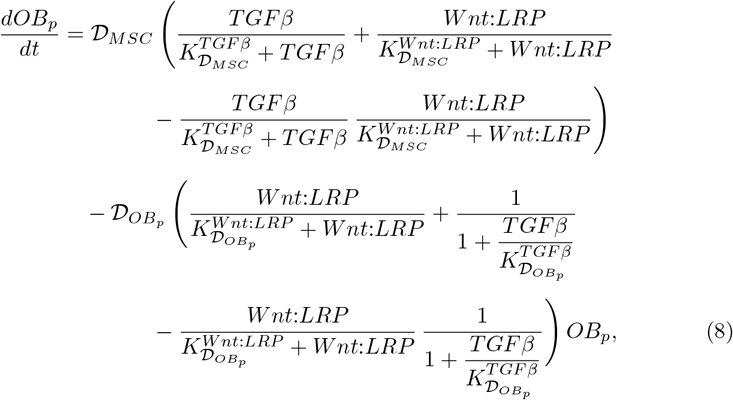

where *TGFβ* is the TGF-*β* concentration; *Wnt*:*LRP* is the concentration of the Wnt-LRP5/6 complex; 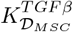 and 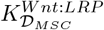 are the corresponding activation coefficients for TGF-*β*- and Wnt signaling-mediated differentiation of MSCs; 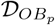 is the maximal maturation rate of osteoblast progenitors; and 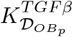 and 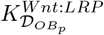 are the repression and activation coefficients for TGF-*β*- and Wnt-mediated maturation of osteoblasts, respectively.

#### Active osteoblasts (*OB*_*a*_)

Active osteoblasts are generated from osteoblast precursors, with their differentiation promoted by Wnt signaling and inhibited by TGF-*β*. At the end of their lifespan, osteoblasts either differentiate into osteocytes or undergo apoptosis.

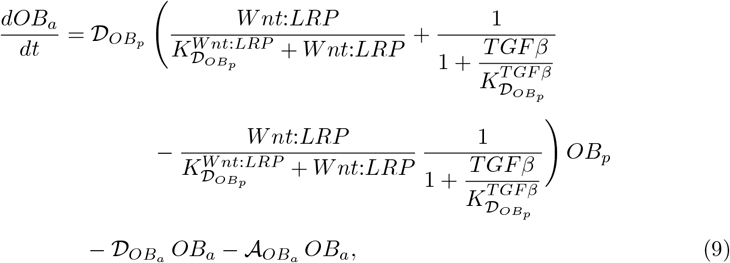

where *OB*_*a*_ is the active osteoclast density; 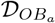 is the osteoblast-to-osteocyte differentiation rate constant; and 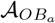 is the osteoblast apoptosis rate constant.

#### Osteocytes (*OT*)

Osteocytes regulate bone homeostasis by producing the Wnt inhibitor sclerostin [36]. They originate from osteoblasts and undergo apoptosis at the end of their long lifespan [37]. Therefore, the population of osteocytes can be described as follows:

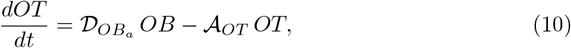

where *OT* is the osteocyte density within the BMU; and 𝒜_*OT*_ is the osteocyte apoptosis rate, assumed to be constant.

#### Receptor activator of nuclear factor *κ*B (*RANK*)

RANK-RANKL-OPG system is the key regulator of osteoclast differentiation. RANK is a type I homotrimeric transmembrane receptor present on osteoclast precursor cells [3, 17]. Binding of RANK to its ligand, RANKL, is required for osteoclast maturation and activation [33, 38, 39]. We assume that all osteoclast progenitor cells express equal amounts of RANK receptors, making total RANK abundance proportional to osteoclast progenitor density, with the proportionality constant equal to the number of RANK receptors per cell, 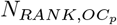. Since we assume that RANK is present on *OC*_*p*_ at a constant density, we omit recycling terms from the equation. At the same time, the degradation term for RANK-RANKL complex is retained to control total RANKL in the system. As a consequence, we must introduce the RANK-RANKL degradation term as an additional source in the RANK equation to maintain a constant RANK density.

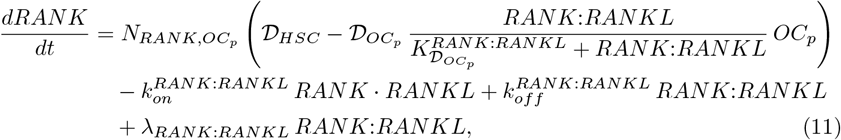

where *RANK* and *RANKL* are the concentrations of RANK and RANKL within the BMU; 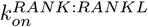 is the association rate constant for RANK binding RANKL; 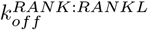 is the dissociation rate constant for RANK-RANKL complex; and *λ*_*RANK*:*RANKL*_ is the degradation rate constant for the RANK-RANKL complex.

#### Receptor activator of nuclear factor *κ*B ligand (*RANKL*)

RANKL is a type II homotrimeric transmembrane protein that is expressed in both membrane-bound and soluble forms [3]. Previous studies have demonstrated that, while soluble RANKL may have a role in maintaining a proper function of osteoclasts, its depletion did not affect the bone loss in estrogen-deficient mice, and membrane-bound RANKL is the form predominantly contributing to bone remodeling [3, 40, 41]. This implies that RANK-RANKL binding must happen via direct cell-cell interaction rather than via paracrine signaling. While all types of osteoblastic lineage cells express RANKL, only osteoblast progenitor cells are likely to be in the appropriate location within the BMU that facilitates receptor binding and osteoclast maturation, as was suggested in the theoretical study by Pivonka et al. (2008) [17]. We adapt this assumption in our model, and consider osteoblast precursors to be the only source of actionable RANKL. RANKL is produced by *OB*_*p*_ with a maximal rate *P*_*RANKL*_ and is eliminated with the degradation rate constant *λ*_*RANKL*_. Expression of RANKL is stimulated by PTH and inhibited by estrogen [17, 32, 42, 43]. Upon its presentation on cell surface, RANKL can either bind to RANK, or to its decoy receptor, OPG [3]. The following equation described RANKL dynamics:

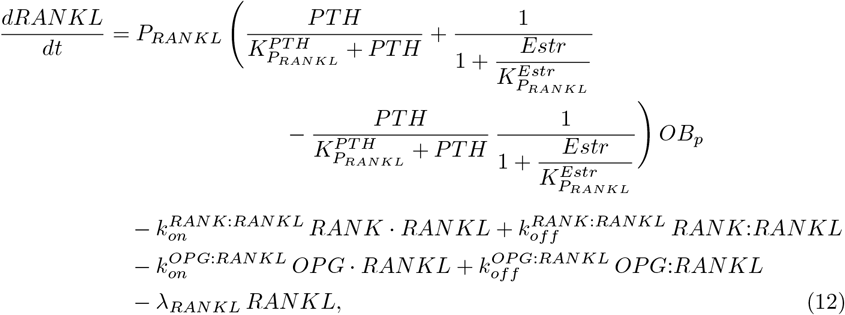

where *PTH* is the parathyroid hormone concentration; 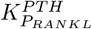 is the activation coefficient for RANKL expression mediated by PTH; 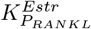 is the repression coefficient governing estrogen-mediated inhibition of RANKL production; and 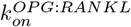 and 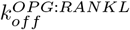 are the association and dissociation rate constants for OPG-RANKL binding, respectively.

Dynamics of RANK-RANKL and OPG-RANKL complexes can be described as follows:

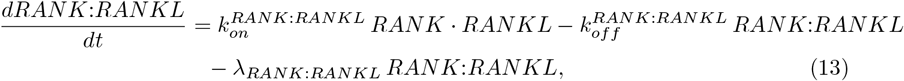

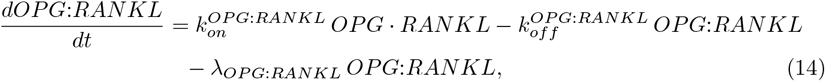

where *OPG* is the OPG concentration within the BMU; and *λ*_*OPG*:*RANKL*_ is the degradation rate constant for OPG-RANKL complex.

#### Osteoprotegerin (*OPG*)

OPG is expressed by osteoblasts and serves as a soluble decoy receptor for RANKL, thereby acting as an endogenous inhibitor of osteoclast differentiation [3]. Generally, OPG expression is regulated by the same factors as RANKL, but in the opposite direction [3]. In line with this trend, PTH has been shown to inhibit OPG expression [44, 45], while estrogen has been demonstrated to enhance its production [12, 46, 47]. In addition, canonical Wnt signaling has been implicated in stimulation of OPG production, independently of its effect on osteoblast differentiation [3, 48, 49].

In their theoretical modeling study, Pivonka and the coleagues (2008) demonstrated that mature osteoblasts constitute the major source of OPG within the BMU [17]. Following that formulation, we assume that OPG is only secreted by *OB*_*a*_ with a maximal production rate *P*_*OPG*_, which is regulated by PTH, estrogen and Wnt signaling. Upon its secretion into the extracellular space, OPG can bind RANKL, or internalize and undergo lysosomal degradation with a degradation rate constant *λ*_*OPG*_.

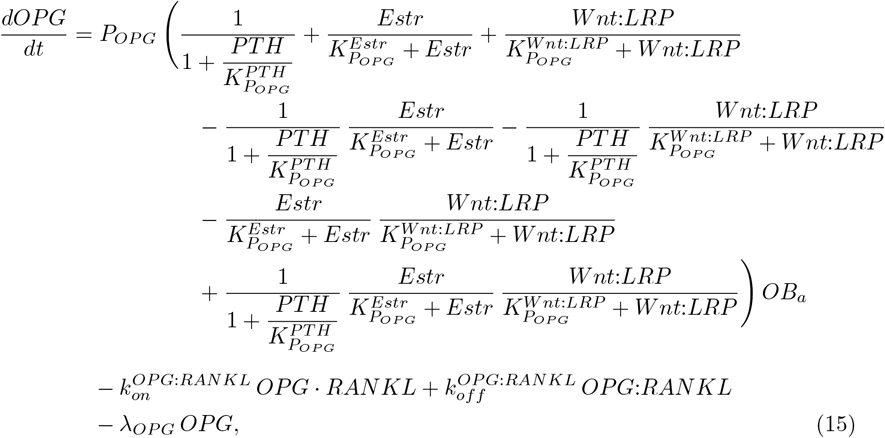

where 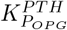 is the represssion coefficient determining PTH-mediated inhibition of OPG secretion; and 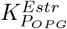 and 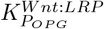 are the activation coefficients for OPG production, mediated by estrogen and Wnt signaling, respectively.

#### Wnt

Wnt proteins comprise a large family of signaling molecules that play an important role in development and are commonly involved in the regulation of cell fate decisions, cell proliferation, migration, polarity, survival and differentiation [50]. There are 19 known structurally related Wnt proteins in humans that either function through the *β*-catenin-dependent canonical pathway or the non-canonical (*β*-catenin-independent) pathway [50]. Although both canonical and *β*-catenin-independent pathways are shown to play a role in bone homeostasis, we only consider canonical Wnt signaling to reduce the model complexity complexity of our model, which is consistent with prior model structures [5, 23, 32, 50]. Canonical Wnt pathway involves binding of Wnt molecules to one of the Frizzled (Fzd) proteins, followed by formation of a ternary complex with the low-density lipoprotein receptor-related protein (LRP) 5/6 [50]. As dysregulation of Wnt-Fzd binding has not been implicated in abnormalities in adult bone remodeling, we do not explicitly model this step, while including LRP5 binding. Wnt is assumed to be secreted into the BMU with a constant production rate 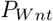 and eliminated with a degradation rate constant *λ*_*Wnt*_. Wnt dynamics is described in our model as follows:

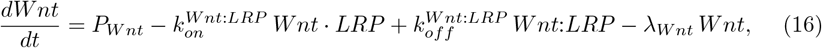

where *Wnt* and *LRP* are the concentrations of actionable Wnt and LRP5/6 in the BMU; and 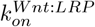 and 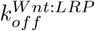 are the association and dissociation rate constants for Wnt-LRP5/6 binding.

#### Dickkopf-1 (*DKK1*)

Dickkopf (DKK) proteins are secreted by osteoblast progenitors and mature osteoblasts and bind to LRP5/6 to inhibit canonical Wnt signaling [32, 50, 51]. Previous studies suggested that the inhibitory action of DKK1 is sufficient to suppress Wnt signaling; therefore, we include DKK1 as the major Wnt inhibitor of the DKK family [52].

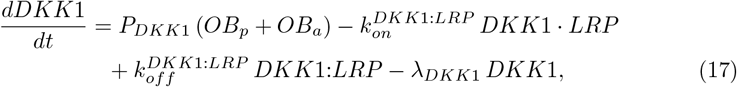

where *DKK*1 is the concentration of DKK1 within the BMU; *P*_*DKK*1_ is the DKK1 secretion rate by osteoblasts and osteoblast progenitor cells; *λ*_*DKK*1_ is the DKK1 degradation rate constant; and 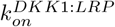 and 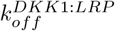 are the association and dissociation rate constants for DKK1-LRP5/6 binding.

#### Sclerostin (*SOST*)

Sclerostin, encoded by *SOST* gene, is a secreted glycoprotein, primarily produced by osteocytes [37, 50, 53]. Sclerostin binds to LRP5/6, thereby inhibiting Wnt signaling [37, 50]. Sclerostin production is negatively regulated by estrogen and PTH [14, 15, 54, 55]. We describe sclerostin dynamics as follows:

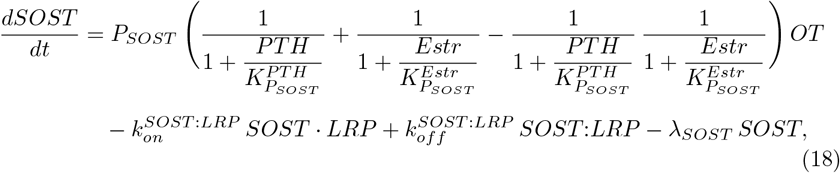

where *P*_*SOST*_ is the maximal production rate of sclerostin by osteocytes; *SOST* is the concentration of sclerostin within the BMU; 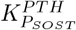 and 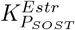 are the repression coefficients for the inhibition of sclerostin production mediated by PTH and estrogen, respectively; 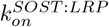 and 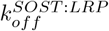 are the association and dissociation rate constants for sclerostin-LRP5/6 binding; and *λ*_*SOST*_ is the sclerostin degradation rate constant.

#### Low density lipoprotein receptor-related protein 5/6 (*LRP*)

Binding of Wnt-Fzd complex to LRP5/6 receptors initiates canonical Wnt signaling. Structural findings suggest that both DKK1 and sclerostin bind primarily to first *β*-propeller (E1) of the LRP5/6 ectodomain, a region required for the formation of the Wnt-Fzd-LRP complex [56, 57]. In addition, functional studies have shown that inhibition of *β*-catenin reporter activity by sclerostin could be reversed by overexpressing LRP5 [58]. Therefore, we treat DKK1 and sclerostin as competitive antagonists and assume that only free LRP5/6 receptors are available for binding. Moreover, we represent LRP5 and LRP6 as a single effective co-receptor pool (LRP5/6), since both are highly homologous (71% homology) Wnt co-receptors and available data do not permit separate identification of LRP5- and LRP6-specific kinetic parameters [59]. We assume that there is a constant density of LRP5/6 receptors on osteoblasts and osteoblast progenitor cells (*N*_*LRP,OB*_), and do not include LRP5/6 production and recycling terms in the equation. Therefore, the free receptor density is governed by osteoblast dynamics and by LRP5/6 binding kinetics. To ensure mass balance for total LRP5/6 in the system, we add additional source terms for LRP5/6 counteracting the degradation of molecular complexes involving the receptor.

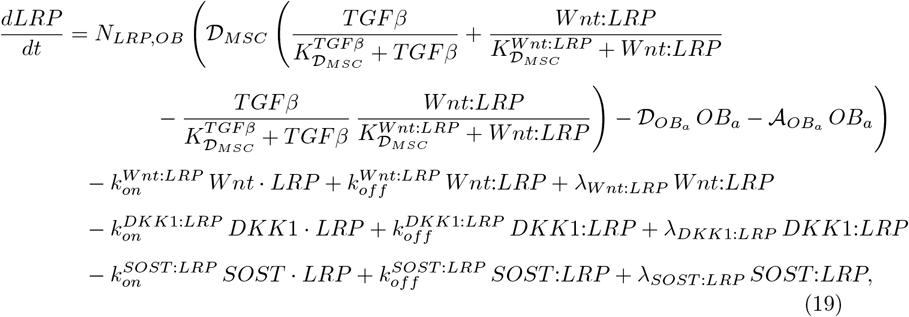

where *λ*_*Wnt*:*LRP*_, *λ*_*DKK*1:*LRP*_, and *λ*_*SOST* :*LRP*_ are the degradation rate constants for the Wnt-LRP5/6, DKK1-LRP5/6, and sclerostin-LRP5/6 complexes, respectively.

The dynamics of the Wnt-LRP5/6, DKK1-LRP5/6, and sclerostin-LRP5/6 complexes can then be described as follows:

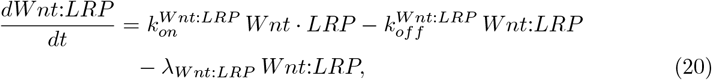

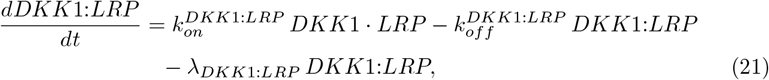

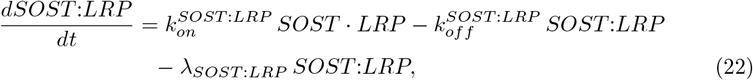

#### Parathyroid hormone (*PTH*)

Parathyroid hormone (PTH) is synthesized by the chief cells of the parathyroid glands [60]. PTH plays an essential role in maintaining Ca^2+^ homeostasis, mainly through its anabolic and catabolic effects on bone [60]. Normally, PTH is present in the bone at steady-state levels. As Ca^2+^ levels drop upon being used during the bone formation, parathyroid gland starts producing PTH [32, 60]. PTH binds to its receptor, PTH1R, on osteoblasts to promote bone resorption via up-regulating RANKL and down-regulating OPG, which results in increased Ca^2+^ release [44, 45, 60]. At the same time, PTH exerts its anabolic effect via inhibiting sclerostin production by osteocytes [54, 60]. We assume that under healthy conditions, PTH is present at steady-state levels and its production equals its degradation. Increase in Ca^2+^ levels, in turn, signals parathyroid gland to stop producing PTH [61]. PTH dynamics can then be described as follows:

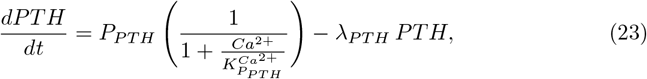

where *P*_*PTH*_ is the PTH production rate in the bone; 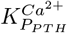 is the repression coefficient describing inhibition of PTH production by Ca^2+^; and *λ*_*PTH*_ is the PTH degradation rate constant.

#### Transforming growth factor *β* (*TGF-β*)

Transforming growth factor-*β* isoforms 1–3 are the growth factors that are stored in the bone matrix in a latent form [32, 62]. During bone resorption, latent TGF-*β* is released from the bone matrix and is subsequently activated through proteolytic processing and conformational changes mediated by osteoclast activity [62, 63]. Active TGF-*β* plays an important role in regulating bone remodeling by modulating osteoblast and osteoclast dynamics. Specifically, TGF-*β* promotes the differentiation of MSCs into osteoblast precursor cells while inhibiting osteoblast maturation [32, 64]. In addition, TGF-*β* promotes osteoclast apoptosis [34]. TGF-*β*1 is the predominant isoform present in bone, and therefore, we explicitly model TGF-*β*1 as the representative TGF-*β* signal regulating bone dynamics [65]. To capture the delay in TGF-*β* signaling after its release from bone matrix, we explicitly model latent TGF-*β* (LTGF-*β*) and activated TGF-*β* as two separate species. In our formulation, the concentration of LTGF-*β* released from bone matrix is directly proportional to the amount of bone resorbed, with the proportionality coefficient *µ*, representing the TGF-*β* concentration stored in one percent of bone matrix.

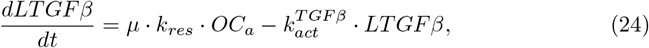

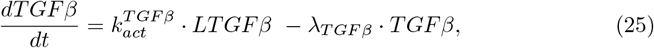

where *LTGFβ* and *TGFβ* are the concentration of TGF-*β* in its latent and active forms, respectively; *k*_*res*_ is the bone resorption rate constant; 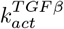 is the TGF-*β* activation rate constant; and *λ*_*TGF β*_ is the TGF-*β* degradation rate constant.

#### Estrogen (*Estr*)

Estrogen plays a key regulatory role in bone remodeling. Estradiol (E2) is the primary biologically active estrogen that mediates estrogen receptor signaling. In postmenopausal women, most circulating E2 originates from peripheral conversion of estrone, which is generated via aromatization of adrenal androgens [66]. In the bone, E2 mediates its effect primarily through estrogen receptor-*α* (ER*α*), which is essential for estrogen-dependent regulation of osteoclast and osteoblast activity, while estrogen receptor-*β* (ER*β*) plays a more limited role [9, 67]. Through ER*α*-mediated signaling, E2 upregulates OPG production, suppresses RANKL and sclerostin expression, and promotes osteoclast apoptosis, exerting a potent anti-resorptive effect [10–15]. To maintain a steady-state E2 concentration in the BMU under baseline conditions, we assume a constant E2 production rate, *P*_*Estr*_ and a first-order degradation rate constant, *λ*_*Estr*_. The equation describing estrogen dynamics takes the following form:

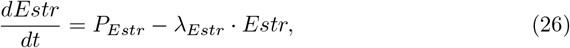

where *Estr* is the E2 concentration within the BMU.

#### Calcium ions (*Ca*^2+^)

Calcium is released from bone matrix upon bone resorption and is incorporated into the bone during formation. As Ca^2+^ becomes depleted during bone formation, its reduced availability limits further mineral deposition, leading to a decrease in bone formation rate. In this way, Ca^2+^ serves as a regulatory control in the model, preventing indefinite bone growth. We model Ca^2+^ dynamics as proposed by Farhat et al (2017), assuming that Ca^2+^ excretion balances daily intake under normal physiological conditions and is inversely regulated by PTH levels [32].

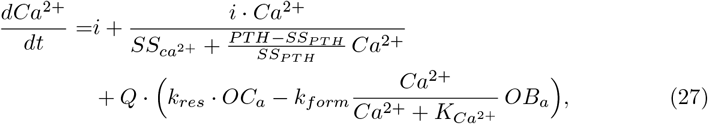

where *Ca*^2+^ is the concentration of Ca^2+^ within the BMU; *i* is the daily Ca^2+^ intake; 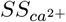 and *SS*_*PTH*_ are the normal steady-state concentrations of Ca^2+^ and PTH, respectively; *Q* is the amount of Ca^2+^ released from one percent of bone; *k*_*form*_ is the bone formation rate constant; and 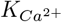 is the concentration of Ca^2+^, at which the rate of bone formation halved.

#### Bone mineral density (*BMD*)

Total bone volume is governed by two opposing processes: bone formation by osteoblasts and bone resorption by osteoclasts. The bone resorption rate is assumed to be directly proportional to the number of active osteoclasts within the BMU. Bone formation depends on both the number of active osteoblasts within the BMU and local calcium availability. Following the approach of Farhat et al., bone formation is modeled as calcium-dependent under low-calcium conditions, while remaining independent of calcium when calcium levels are sufficient [32]. Changes in bone mineral density (BMD) are determined by the net balance between bone formation and resorption over time and by the bone mineral content, which is assumed to remain constant in the model [22].

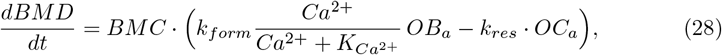

where *BMD* is in the percent unit (assumed to be 0 at baseline); and *BMC* is the bone mineral content.

#### Bone turnover metabolites (*BTMs*)

Bone turnover metabolites (BTMs) serve as biomarkers for osteoblast and osteoclast activity that can be measured in blood and urine [68]. Osteoblast-mediated osteoid production and mineralization are associated with the release of bone-specific alkaline phosphatase (BSAP), osteocalcin, and procollagen type I N-terminal propeptide (P1NP), which are commonly used as markers of bone formation [69]. Conversely, osteoclast activity is reflected by the production of N- and C-terminal telopeptides of type I collagen (NTX and CTX), as well as tartrate-resistant acid phosphatase 5b (TRACP5b), which serve as markers of bone resorption [69]. Despite their utility as markers of bone formation and resorption, BTMs exhibit substantial intra- and inter-individual variability, influenced by factors such as circadian rhythm, food intake, age, renal clearance, lifestyle, and analytical assay [68, 69].

Systemic BTM concentrations depend on both their release during bone remodeling and their elimination from blood and urine. While a mechanistic representation would explicitly model biomarker production and elimination kinetics, we adopt a simplified approach in which formation and resorption rates are linked to corresponding BTM levels through proportionality coefficients. In this study, we model BSAP, P1NP, CTX, and NTX, as these biomarkers were reported in the intervention studies used for model calibration and sensitivity analysis. Given the weak correlation between BTMs and BMD, as well as substantial variability in BTM levels, we focus on reproducing qualitative, rather than quantitative, changes in biomarkers in response to interventions.

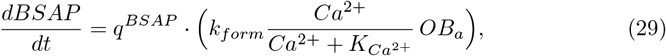

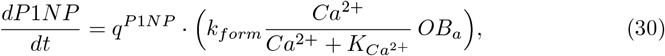

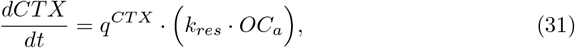

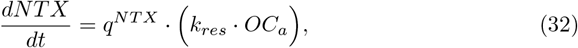

where *q*^*BSAP*^, *q*^*P* 1*NP*^, *q*^*CTX*^, and *q*^*NTX*^ are the proportionality coefficients relating bone formation and resorption rates to the respective biomarkers.

### Model calibration

#### Parameter identification

The base model contains 27 ordinary differential equations and 64 parameters. For initial parameterization of the model, we conducted an extensive literature review. Cellular differentiation and apoptosis rates, molecule secretion rates and half-lives, and activation and repression coefficients were extracted from *in vivo* and *in vitro* studies when available, and from previously published mathematical models otherwise. Cell densities were calculated based on assumptions about total cell counts in the body for the specific cell type and the total number of BMUs in the body, similarly to previously used approaches [32, 70]. Initial conditions for molecular concentrations and daily Ca^2+^ intake were extracted from clinical reference values in postmenopausal women. The amount of Ca^2+^ and TGF-*β* stored in a bone unit, BMC, and bone formation and resorption rate constants were adapted from previous models of bone remodeling. Table 1 summarizes the values of the 50 parameters used in the model that were fixed based on literature review.

**Table 1.**
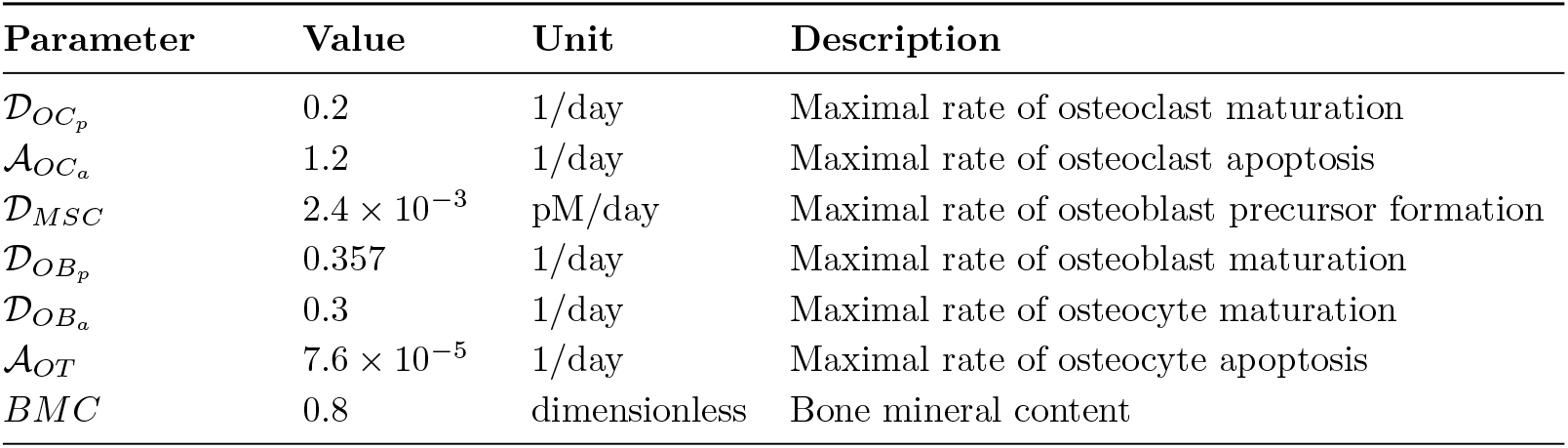
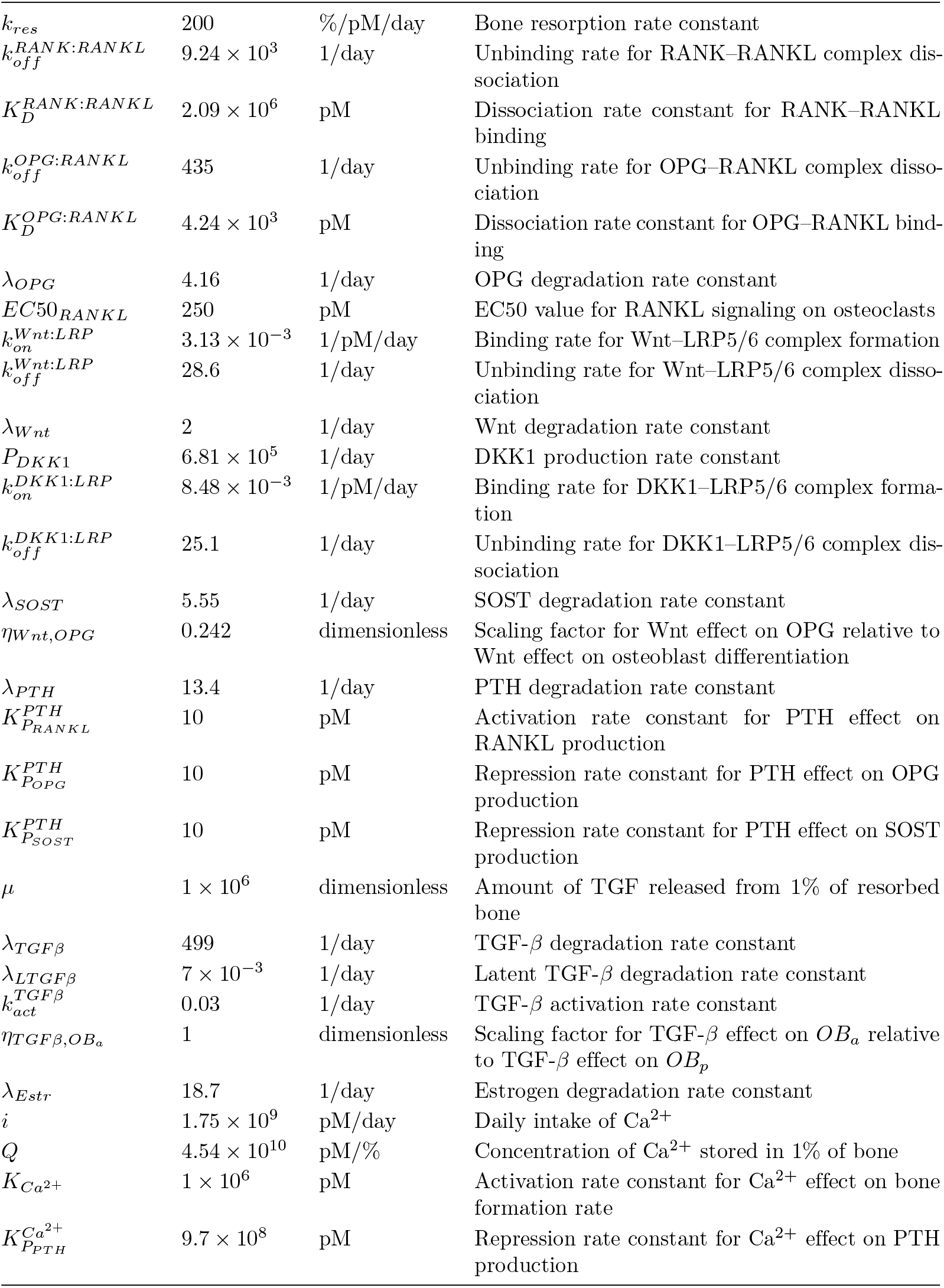
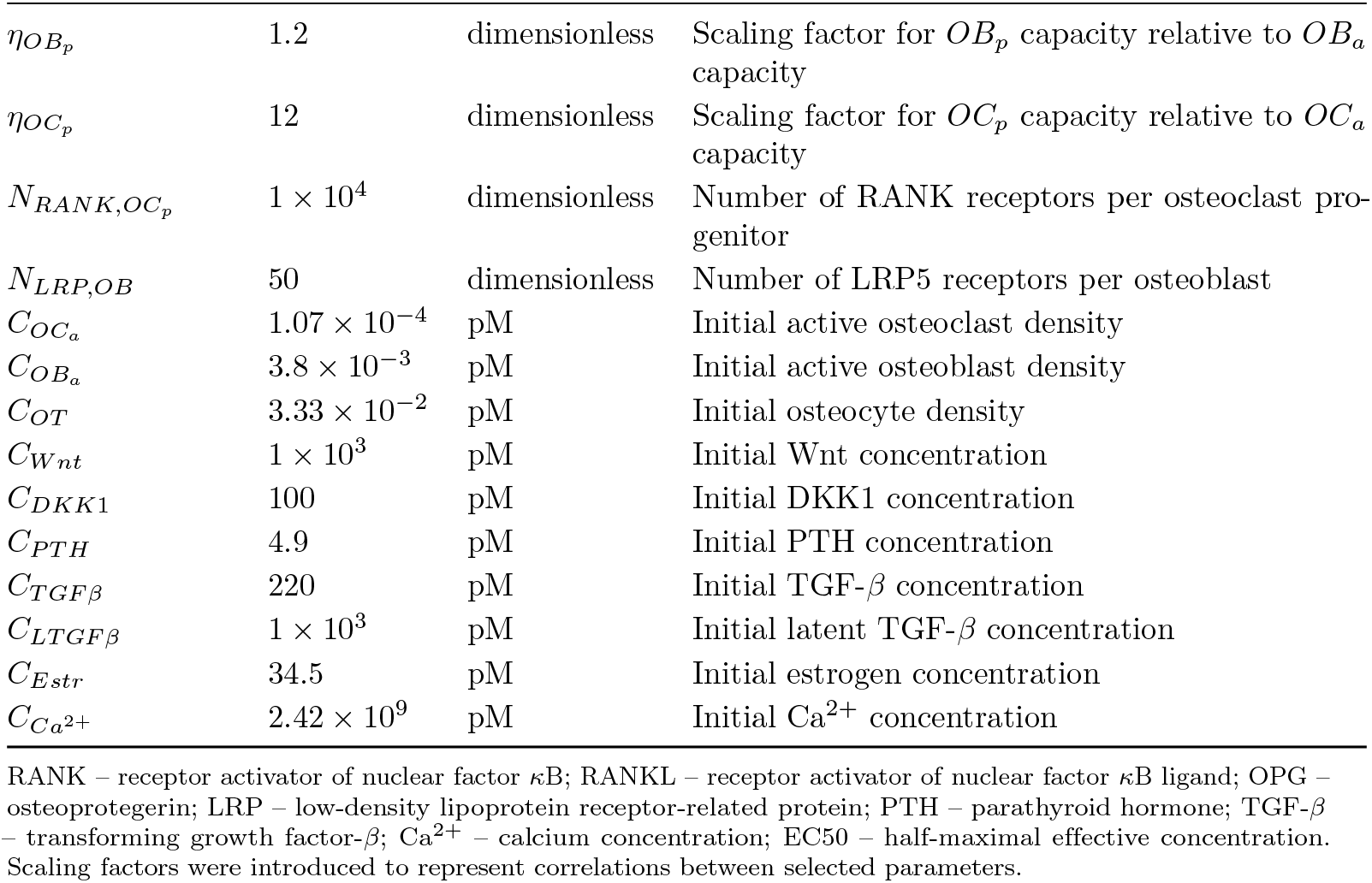
Fixed parameters describing the bone remodeling model.

Upon initial parameterization of the model, we conducted sensitivity analysis for the parameters with the higher degree of uncertainty and selected 14 parameters for calibration to clinical data. The calibration aimed to capture healthy bone dynamics in postmenopausal women and the effects of bone-modifying interventions, such as denosumab, romosozumab, and micronized estradiol [71–74].

#### Calibration to clinical data

To calibrate the healthy BMD dynamics in postmenopausal women, we digitized the data on volumetric BMD (vBMD) of cortical bone at the distal radius from the prior population study of age-related bone loss in humans [71]. Since we model BMD loss as the percent loss from baseline (menopause onset), the extracted vBMD in mg/cm^3^ were converted to percent loss units. To establish vBMD at baseline, we assumed 50 years to be the average age of menopause onset, and linearly extrapolated the data to obtain the estimate for population mean at that age [75, 76]. The extrapolated vBMD value at baseline was set to 0 percent, and all subsequent values were converted to percent change from baseline.

To construct the hybrid datasets for BMD change following therapeutic interventions, we obtained the average age of participants enrolled in each trial and simulated healthy postmenopausal BMD dynamics for the duration between menopause onset and treatment initiation. Changes in BMD following the intervention that were reported in the trial were then calculated relative to the simulated BMD value at treament initiaion. To model BTM dynamics following the intervention, we assumed constant baseline BTM levels between menopause onset and treatment initiation, and calculated percent changes in BTMs from baseline during the course of treatment, as reported in the corresponding clinical trials. Proportionality coefficients relating bone formation and resorption rates to the respective biomarkers were calculated to match the baseline values. All clinical data were digitized using WebPlotDigitizer (version 5.2) [77].

As different interventions perturbed separate regulatory components of the system and produced unique patterns in BTM dynamics, each intervention enabled identifying a unique set of parameters, which were estimated by sequentially fitting the model to three intervention-specific datasets. Each dataset comprised trajectories for BMD, a bone formation marker (BSAP or P1NP), and a bone resorption marker (CTX or NTX). To prioritize accurate reproduction of BMD trajectories, post-intervention BMD data were weighted 10-fold higher than BTM data during model fitting. In addition, to enforce a stricter fit to healthy BMD decline, pre-intervention BMD data points were weighted 10-fold higher than post-intervention BMD data. Parameter estimation was performed using the Particle Swarm Optimization algorithm in MATLAB Global Optimization Toolbox (Version 24.1), followed by local optimization using the Nelder-Mead simplex algorithm [78, 79]. Model parameters were estimated by minimizing a weighted sum of squared residuals between model predictions and observed BMD and BTM trajectories, assuming a constant error model. During each objective function evaluation, the model was simulated to steady state to ensure equilibration.

### PKPD modeling of interventions

To calibrate the BMD dynamics following treatment with bone-modifying agents, we deemed it important to accurately represent the perturbations induced by the intervention on the mechanistic level. Due to their well-defined mechanisms of action, we selected three interventions for model calibration: the RANKL inhibitor denosumab, the sclerostin inhibitor romosozumab, and hormonal regulator micronized E2.

#### Denosumab and romosozumab models

Denosumab is a fully human recombinant monoclonal antibody that binds to RANKL, inhibiting osteoclast maturation and slowing down bone resorption [80]. Romosozumab is a humanized monoclonal antibody that binds to sclerostin, thereby enhancing Wnt signaling, which produces an anabolic effect on bone [81]. Both antibodies are administered subcutaneously (SC), and exhibit non-linear pharmacokinetics (PK) characterized by a decrease in apparent clearance with increasing dose, suggesting target-mediated drug disposition (TMDD) [81, 82]. The PK of denosumab and romosozumab were described using a minimal physiologically based pharmacokinetic (mPBPK) model for antibody distribution, with an explicit bone compartment to enable estimation of antibody concentrations in bone (Figure 2) [83]

**Fig 2.**
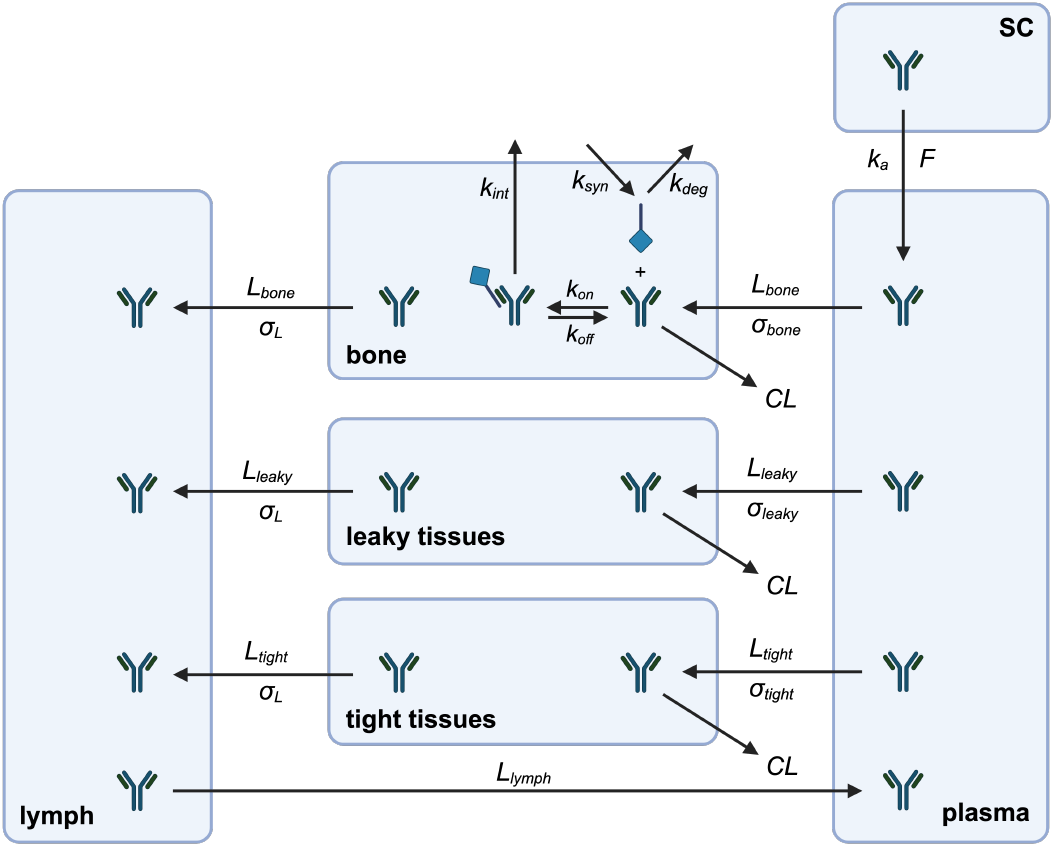
Minimal PBPK model for denosumab and romosozumab distribution. The model describes the distribution of modeled antibodies within the body. The subcutaneously (SC) administered drug is absorbed into plasma and is distributed into tight (skin, adipose, muscle, brain), leaky (kidney, heart, etc.) tissues, and bone. The lymph flow from the tissues returns the drug back to the plasma. Antibodies are cleared from the body by non-specific internalization and lysosomal degradation in the tissues or by target-mediated drug disposition (TMDD) in the bone. TMDD was implemented as a full TMDD model. *CL – non-specific clearance; k*_*a*_ *– absorption rate constant; F – apparent bioavailability; L – flow; σ – vascular reflection coefficient; k*_*syn*_ *– target synthesis rate; k*_*deg*_ *– target degradation rate constant; k*_*on*_ *and k*_*off*_ *– antibody-target association and dissociation rate constants; k*_*int*_ *– antibody-target internalization rate constant*.

In short, the antibody is administered in an SC compartment (*V*_*sc*_), from which it is absorbed into the plasma compartment (*V*_*p*_). From plasma, antibody travels to tight (*V*_*tight*_, adipose, skin, muscle and brain), leaky (*V*_*leaky*_, heart, liver, kidney, etc.) tissues, or bone (*V*_*bone*_) via lymph flow (*L*). Once in tissue, the antibody can undergo non-specific internalization inside the cell followed by lysosomal degradation, modeled as a first-order process with rate constant *k*_*e*_. Alternatively, the antibody can enter the lymph (*V*_*L*_) and return to *V*_*p*_ with the lymph flow. Assuming that pharmacologically relevant RANKL and sclerostin activity is localized to bone tissue, we implemented a full TMDD model within the bone compartment to avoid over-parameterization and identifiability issues. The resulting system of equations describing the mPBPK model is given below.

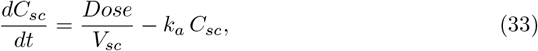

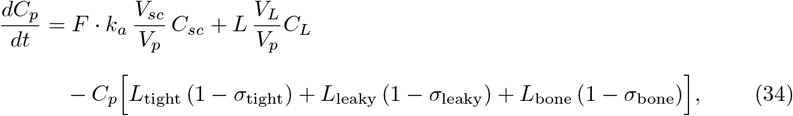

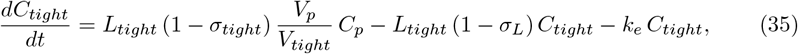

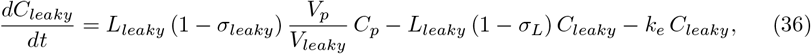

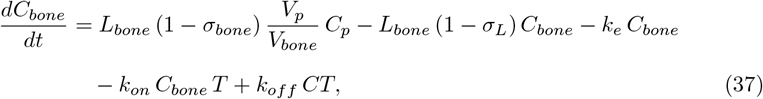

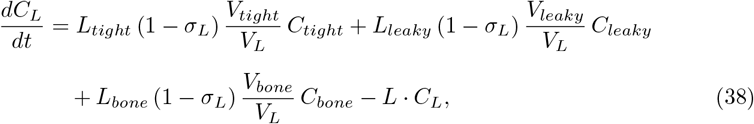

where *C*_*sc*_, *C*_*p*_, *C*_*tight*_, *C*_*leaky*_, *C*_*bone*_, and *C*_*L*_ denote antibody concentrations in the subcutaneous injection site, plasma, tight tissues, leaky tissues, bone, and lymph compartments, respectively; *Dose* is the administered subcutaneous dose, *F* is bioavailability, and *k*_*a*_ is the first-order absorption rate constant from the subcutaneous compartment; *V*_*sc*_, *V*_*p*_, *V*_*tight*_, *V*_*leaky*_, *V*_*bone*_, and *V*_*L*_ denote the volumes of the corresponding compartments; *L*_*tight*_, *L*_*leaky*_, and *L*_*bone*_ represent flow rates from plasma to tight, leaky, and bone compartments, respectively, and *L* denotes return flow to plasma; *σ*_*tight*_, *σ*_*leaky*_, *σ*_*bone*_, and *σ*_*L*_ are vascular reflection coefficients governing antibody extravasation and lymphatic return; *k*_*e*_ is the non-specific elimination rate constant; *T* and *CT* denote free target and antibody–target complex concentrations in bone, while *k*_*on*_ and *k*_*off*_ are the association and dissociation rate constants for antibody binding its respective target (RANKL or sclerostin).

The dynamics of the free target *T* are described by the corresponding equations for RANKL and sclerostin, introduced earlier (Eqs. 12 and 18). The dynamics of the antibody-target complex (*CT*) in bone are given by

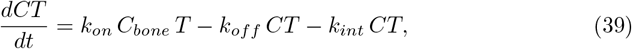

where *k*_*int*_ is the internalization rate constant of the antibody-target complex.

To parameterize the mPBPK model, we used human physiological values for compartment volumes, flows and reflection coefficients [83, 84]. The absolute bioavailabilies of denosumab and romosozumab following SC administration were obtained from literature [85, 86]. For the membrane-bound target RANKL, *k*_*int*_ of the denosumab–RANKL complex was assumed to be equal to that of endogenous RANKL. This assumption links internalization of the denosumab-RANKL complex to physiological RANKL turnover and therefore enables estimation of the degradation rate of free RANKL within the TMDD framework. In contrast, we did not make a similar assumption for the romosozumab-sclerostin complex, as sclerostin is a secreted protein that undergoes renal clearance [87]. The parameters *V*_*p*_, *k*_*a*_, *k*_*e*_, *k*_*int*_ and target capacity were calibrated to fit clinical data.

Denosumab PK profiles following single-dose SC administration in healthy postmenopausal women were obtained and digitized from the FDA Clinical Pharmacology Review (Study 20010124) [88]. The data included concentration-time profiles from 20 postmenopausal women across six dose levels (0.01, 0.03, 0.1, 0.3, 1, and 3 mg/kg).

Romosozumab PK data were obtained from the FDA Multi-Discipline Review (Study 20060220) and included concentration-time profiles following single-dose SC administration in a combined cohort of healthy postmenopausal women and healthy men (n = 42) across six dose levels (0.1, 0.3, 1, 3, 5, 10 mg/kg) [86]. Three dose levels were used for model calibration (0.01, 0.1, and 1 mg/kg for denosumab; 0.1, 1, 5 mg/kg for romosozumab), while the remaining data were reserved for model validation. Parameter estimation was performed using the Particle Swarm Optimization algorithm in MATLAB Global Optimization Toolbox (Version 24.1) assuming an exponential error model, followed by local optimization using the Nelder-Mead simplex algorithm [78, 79]. Minimal PBPK model fit was assessed by the visual predictive check (VPC). In addition, we calculated pharmacokinetic parameters, such as maximum plasma concentration *(C*_*max*_), time to reach maximum concentration *(T*_*max*_), terminal half-life *(T*_*1/2*_) and area under the concentration time curve *(AUC)* for the clinically administered doses of both drugs (60 mg SC for denosumab, 210 mg SC for romosozumab), and compared them to the values reported in the FDA label [89, 90].

#### Micronized estradiol model

Whereas denosumab and romosozumab are administered once every six months (Q6M) and once monthly (QM), respectively, micronized estradiol is administered daily, resulting in a sustained increase in steady-state estradiol concentration over years of treatment [74, 91]. Accordingly, based on data from Prestwood et al. (2003), the effect of daily oral administration of 0.25 mg of micronized estradiol was modeled as a constant shift in steady-state estradiol concentration from the baseline value of 34.5 pM to 95.8 pM, as reported in Prestwood et al. (2003), without explicitly representing PK details [74].

### Mechanism of estrogen regulation

Our structural model assumes that estrogen regulates bone remodeling via four mechanisms: downregulation of RANKL and sclerostin, upregulation of OPG, and promotion of osteoclast apoptosis. To better understand the contribution of each regulation mechanism, we performed an ablation experiment, simulating BMD dynamics under four scenarios: (1) no effect on osteoclast apoptosis; (2) no effect on RANKL production; (3) no effect on OPG production; and (4) no effect on sclerostin production. Annual BMD loss rate was calculated for each of the scenarios and compared with the BMD loss rate under a baseline scenario, which included all four regulatory mechanisms. The experiment was conducted under assumption of normal postmenopausal estrogen levels, as well as under the 90% estrogen suppression scenario.

### Genetic effect incorporation

#### Genetic variant selection

To select genetic variants for inclusion in the model, we performed a literature review of candidate gene studies and genome-wide association studies (GWAS) to identify common single nucleotide polymorphisms (SNPs) with replicated associations with BMD [29, 30, 92–113]. The variants were further restricted to genes whose functional effect could be explicitly represented within the mechanistic model and to those with global minor allele frequency (MAF) ≥ 5%. Global and ancestry-specific MAFs were obtained from the 1000 Genomes Project (genome build GRCh37) and were consistent with the frequencies reported in gnomAD [114, 115]. Reference and variant alleles were assigned based on the Ensembl Genome Browser, which was also used to determine the location and functional consequence of the variant [116]. The final list included 22 SNPs across 8 genes and is presented in Table 2.

**Table 2.**
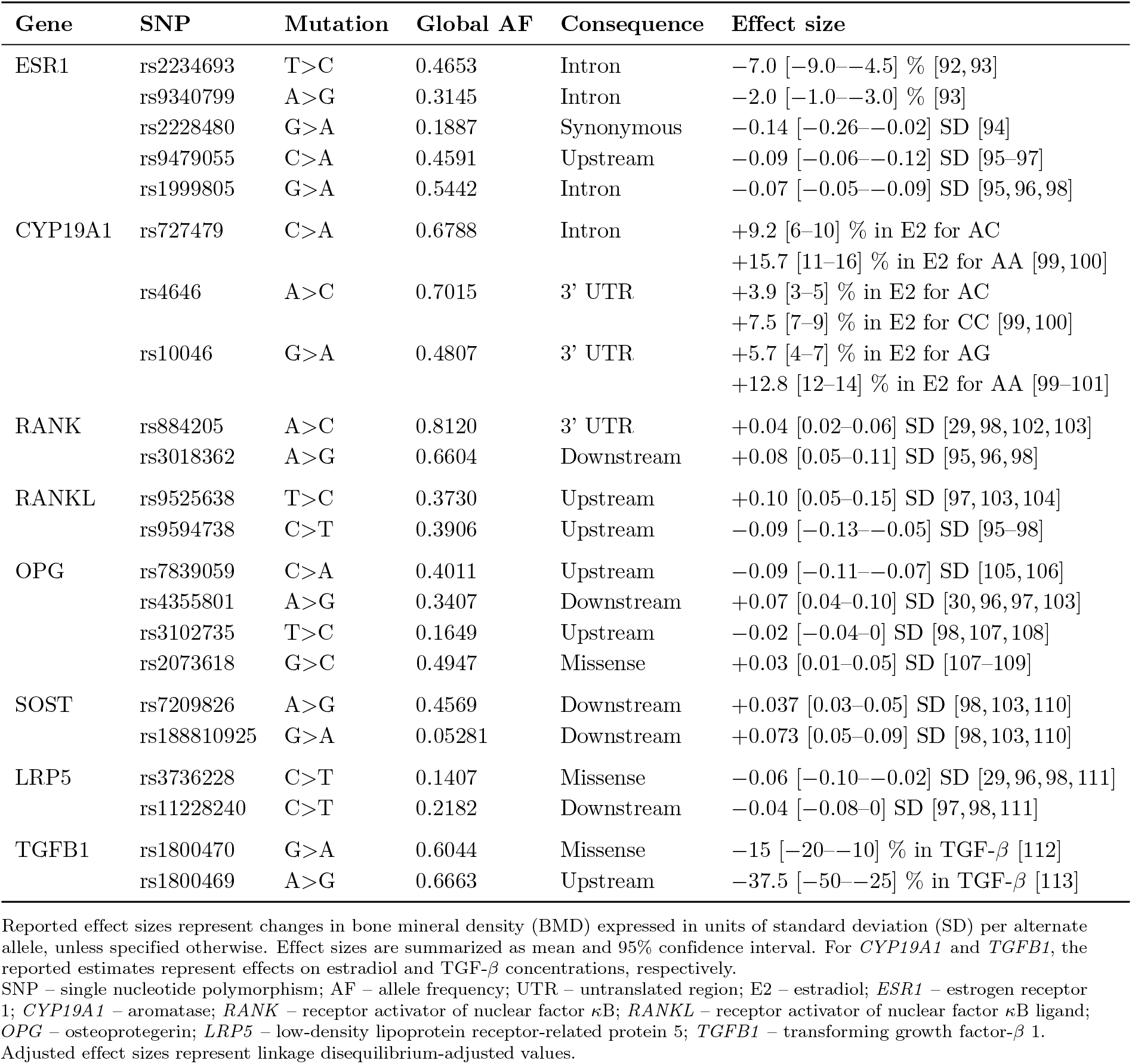
Summary of included SNPs.

Global and ancestry-specific pairwise linkage disequilibrium (LD) measures (*r*^2^, *D*^*′*^) and genotype counts were obtained for SNP pairs located on the same chromosome using LDlink, based on the data from 1000 Genomes Project (GRCh37) [114, 117]. LDlink does not provide pairwise LD metrics for non-biallelic variants. Accordingly, for all LD analyses, *OPG* rs4355801 was represented by a biallelic surrogate SNP in perfect LD with rs4355801 (*OPG* rs10101385; *r*^2^ = 1 across populations), as reported in the Ensembl Genome Browser [116].

#### Mathematical formulation

Genetic effects were mechanistically incorporated into the model by mapping SNPs to their predicted functional effects on molecular processes represented in the model. Based on variant annotation, SNPs were classified as affecting either protein expression (e.g., variants located in regulatory elements, such as promoters, enhancers, untranslated regions or introns) or protein binding properties (e.g., variants located in coding regions).

For genes encoding molecular species explicitly represented in the model (*RANK, RANKL, OPG, LRP5, SOST*, and *TGFB1*), genetic effects on expression were implemented as modifiers of the corresponding production rate constants, whereas variants predicted to affect binding properties were implemented as modifiers of association rate constants (*k*_*on*_). Variants in *CYP19A1*, which encodes aromatase, were incorporated as modifiers of the estrogen production term. Variants in *ESR1*, encoding ER*α*, were incorporated by changing estrogen signaling sensitivity. Regulatory effects are included in the model using Hill-type equations, in which estrogen signaling is represented by activation or repression constants (*K*_*act*_, and *K*_*rep*_), corresponding to E2 concentrations producing half-maximal response. Under this formulation, increased receptor abundance results in enhanced signaling sensitivity, reflected by a reduced apparent *K*. Accordingly, *ESR1* variants affecting receptor expression were modeled as inverse modifiers of *K*_*act*_ and *K*_*rep*_ for estrogen regulation.

An additive genetic model was assumed for all SNPs. For each variant, the marginal effect per alternate allele was represented as a fold change in the affected parameter. Thus, for a given SNP with *n*_*alt*_ alternate alleles and effect size *β*, the affected parameter was adjusted by a genetic modifier *R*, calculated as follows:

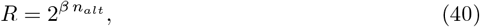

This formulation allows a unified representation of genetic effects across model parameters, such that *P*_*SNP*_ = *P R, k*_*on,SNP*_ = *k*_*on*_ · *R*, and *K*_*SNP*_ = *K/R*.

Due to LD, genetic variants within the same locus are not independent from one another, and marginal effect estimates partially capture shared genetic signal. To avoid double-counting effects of correlated SNPs, marginal per-allele effect estimates were adjusted for LD structure between variants within the same gene, producing conditional estimates used for the calculation of the combined genetic effect [118, 119].

Let 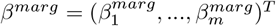 be the vector of marginal effect sizes for *m* standardized genotypes *G* = (*G*_1_, …, *G*_*m*_)^*T*^ . Let **C** denote the genotype correlation matrix with elements *C*_*ij*_ = *corr*(*G*_*i*_, *G*_*j*_). The relationship between marginal and conditional effect sizes 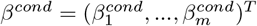 is given by

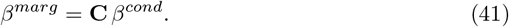

Solving for the conditional effects, we get

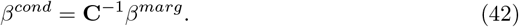

The LD-adjusted estimates represent the effect of each variant conditional on all other variants within the locus and were used to calculate the combined genetic effect. For a given gene, the combined genetic effect was defined as

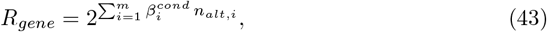

where *n*_*alt,i*_ is the reference allele count for variant *i*, and 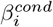 is the LD-adjusted per-allele effect of SNP *i*. In the absence of LD, the correlation matrix reduces to identity matrix and conditional effect sizes equal marginal estimates. As LD increases, conditional effect sizes are attenuated, reflecting partitioning of genetic signal across correlated SNPs.

#### Effect size determination

Marginal per-SNP effect sizes (mean and 95% confidence interval) on total hip BMD were extracted from literature for variants in *ESR1, RANK, RANKL, OPG, SOST*, and *LRP5*. For *CYP19A1* and *TGFB1*, the extracted per-SNP effects were associated with circulating E2 and TGF-*β* levels, respectively. Reported effect sizes were translated into expected percent changes in BMD or shift in the corresponding molecular concentrations at 5 years following menopause onset. We assumed that genetic effects remain consistent over time. Therefore, a reported per-allele effect of 0.10 standard deviations (SD) in BMD was interpreted as an increase in the rate of BMD loss, such that each alternate allele increased cumulative BMD loss over 5 years by 0.10 SD. We assumed a standard deviation of 2% for BMD loss over 5 years following menopause onset, based on the range of values reported in the literature [120–127].

The base model was used to simulate the expected 5-year BMD decline for the reference genotype, defined as the genotype with the highest expected frequency based on global allele frequencies. Expected BMD trajectories for the remaining genotypes were then computed as deviations from the baseline simulation according to the per-allele effect size. The genotype-specific BMD values were subsequently used for estimating the corresponding mechanistic effect parameters (*β*) independently for each SNP. Parameter optimization was performed using the Particle Swarm Optimization algorithm with constant error. To propagate uncertainty in the reported effect size, we repeated optimization 100 times, randomly sampling the expected BMD change from a normal distribution defined by the published mean and standard error inferred from the 95% CI, assuming large-sample normality. The resulting distributions represent marginal *β* estimates and were used as inputs for subsequent simulations.

### Population modeling

To evaluate the combined effect of multiple genetic perturbations, we simulated BMD dynamics in a diverse virtual population of 1,000 individuals. The population was stratified by ancestry in a 60–25–15 proportion corresponding to European (EUR), African (AFR), and East Asian (EAS) ancestries, respectively. Within each ancestry group, individual genotypes were randomly sampled from a joint genotype distribution that preserved the ancestry-specific allele frequencies and LD patterns between variants. Pairwise correlations between SNPs located on the same chromosome, as reported in the 1,000 Genomes Project, were used to construct ancestry-specific covariance matrices that defined the joint genotype distributions. BMD dynamics was simulated in the virtual population over a 5-year period. For the virtual population analysis, we defined the primary outcome as the percent BMD loss at 5 years.

#### Model assessment

For the population-level validation, we obtained BMD loss data from previously published clinical trials and observational studies [120–127]. All of the included studies reported longitudinal BMD loss. When available, the exact values were extracted from table or text and digitized from the figures otherwise. We then calculated mean, standard deviation, median, interquartile range (IQR), and 95% prediction interval for 5-year BMD loss and compared the simulated distribution with the literature-reported values. Fraction of variability attributable to genetics was determined by variance decomposition with the assumed total variability in 5-year BMD loss of 2% (the value used to estimate genetic effects). To separate inter-individual variability in genetics from uncertainty in genetic effect estimates, we simulated BMD dynamics under two scenarios: (i) using fixed genetic effects for each SNP; and (ii) incorporating uncertainty in genetic effect estimates. The total variability can then be decomposed as

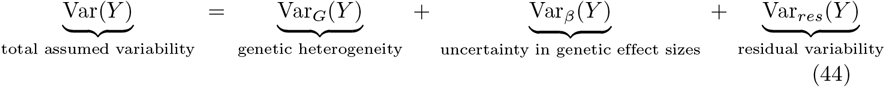

The first simulation scenario, under fixed genetic effects, allows us to estimate Var_*G*_(*Y*), while the scenario with incorporated uncertainty in genetic effect estimates gives us the combined Var_*G*_(*Y*) + Var_*β*_(*Y*).

#### Subgroup analysis

To further characterize variability in BMD loss within the virtual population, we stratified the analysis by genetic ancestry. For each ancestry group (EUR, AFR, and EAS), we evaluated the distribution of 5-year BMD loss using the median, IQR, and 95% prediction interval. Statistical differences between ancestry-specific distributions of BMD loss were assessed using pairwise Wilcoxon rank-sum test. A similar analysis was conducted at the genotype level for each SNP, where the outcomes were calculated for each genotype and compared using the Wilcoxon rank-sum test. P-values were adjusted for multiple testing using Bonferroni correction.

#### Genetic interaction analysis

To better understand how SNP-SNP interactions affected BMD dynamics in the virtual population, we fit an additive genetic model with an interaction term for all pairwise SNP combinations [128]. For each pair of SNPs, predicted BMD loss was modeled as

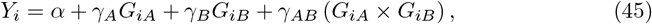

where *Y*_*i*_ denotes the simulated BMD loss for individual *i, G*_*iA*_ and *G*_*iB*_ are alternate allele dosages for the two variants, *α* is the intercept corresponding to the expected BMD loss for individuals with zero alternate alleles at both loci, *γ*_*A*_ and *γ*_*B*_ represent the additive genetic effects of variants *A* and *B*; and *γ*_*AB*_ quantifies the deviation from additivity in the joint genetic effect.

Positive *γ*_*AB*_ values indicate a synergistic effect of the two SNPs, whereas negative values suggest attenuation of additive effects. Statistical significance was assessed with two-sided Wald test. P-values were adjusted for multiple testing using the Bonferroni correction.

## Results

### PKPD models for therapeutic interventions

Distribution of denosumab and romosozumab into tissues was described using mPBPK models incorporating SC absorption, non-specific clearance from tight, leaky and bone tissues, and TMDD in the bone tissue. Separate models were developed and parameterized for each therapeutic agent.

The denosumab PK model was calibrated using clinical concentration-time data after a single-dose SC administration of 0.01, 0.1, and 1 mg/kg of denosumab, and validated using 0.03, 0.3, and 3 mg/kg doses (Fig 3A). The calibrated model accurately captured nonlinear PK of denosumab over the dose range. At lower doses, rapid target binding and internalization dominated drug elimination, whereas at higher doses, saturation of RANKL resulted in linear clearance.

**Fig 3.**
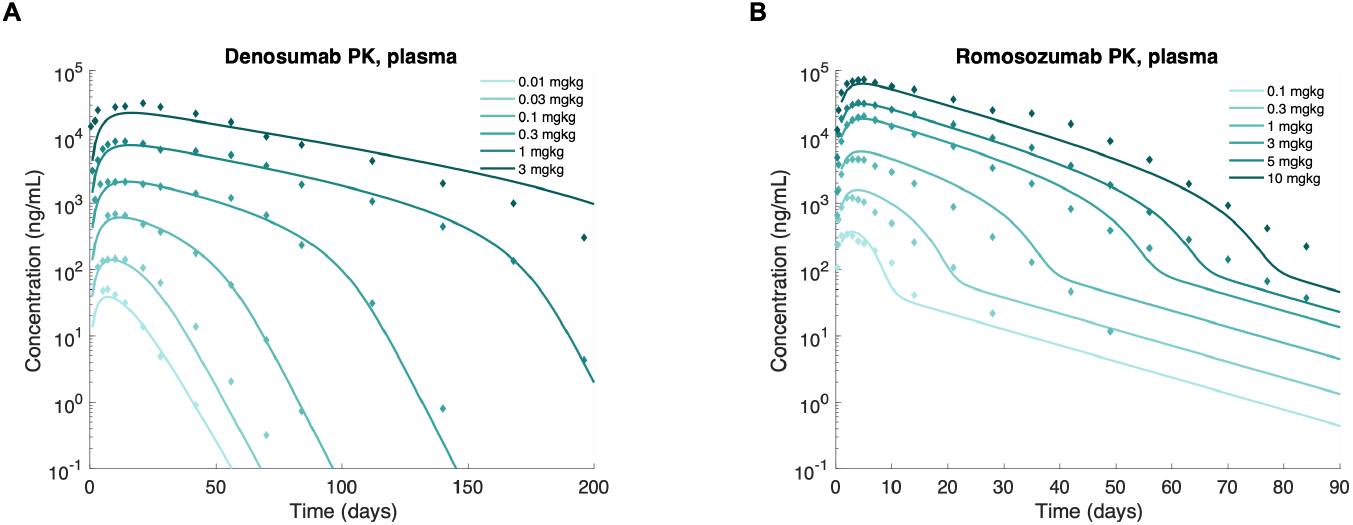
Denosumab and romosozumab PK. Minimal physiology-based pharmacokinetic (PBPK) model fits for (A) single dose subcutaneous (SC) administration of 0.01–3 mg/kg of denosumab and (B) single dose SC administration of 0.1–10 mg/kg of romosozumab. The denosumab model was calibrated on 0.01, 0.1, and 1 mg/kg, and validated on 0.03, 0.3, and 3 mg/kg data. The romosozumab model was calibrated on 0.1, 1, 5 mg/kg, and validated on 0.3, 3, 10 mg/kg data.

An analogous mPBPK model for romosozumab was calibrated using clinical data for single dose SC administration of 0.1, 1, and 5 mg/kg of romosozumab, with validation performed at 0.3, 3, and 10 mg/kg (Fig 3B). The model captured observed plasma concentration-time profiles across all six doses. In contrast to denosumab, romosozumab exhibited more complex clearance behavior characterized by three apparent slopes in the elimination phase, due to rapid dissociation of the romosozumab-sclerostin complex (*k*_*off*_ = 2.51 *×* 10^1^ day^-1^), which was substantially faster than the degradation of the complex (*k*_*int*_ = 5.59 *×* 10^−2^ day^-1^).

Estimated PK parameters for denosumab and romosozumab (*V*_*p*_, *k*_*a*_, *k*_*e*_, *k*_*int*_, *C*_*RANKL*_, and *C*_*SOST*_) are summarized in Table 3. For the clinically administered doses of denosumab (60 mg SC) and romosozumab (210 mg SC), the model-predicted exposure metrics *(C*_*max*_, *T*_*max*_, *T*_*1/2*_, *AUC)* were consistent with values reported in the FDA label (Table 4). Overall, the mPBPK models accurately described denosumab and romosozumab PK across a wide range of doses, supporting their use for model calibration based on the pharmacodynamic (PD) response to these medications.

**Table 3.** Minimal PBPK model parameters.

| Parameter | Value | Unit | Description | Source |
| --- | --- | --- | --- | --- |
| *Physiological parameters* |  |  |  |  |
| $V_{tight}$ | $1.07 \times 10^{-1}$ | L | Tight tissue volume (skin, adipose, muscle, brain) | [84] |
| $V_{leaky}$ | $2.65 \times 10^{-2}$ | L | Leaky tissue volume (heart, liver, kidney, etc.) | [84] |
| $V_{bone}$ | $2.66 \times 10^{-2}$ | L | Bone tissue volume | [84] |
| $V_L$ | $3.86 \times 10^{-3}$ | L | Lymph volume | [84] |
| $L$ | 2.90 | L/day | Total lymph flow | [83] |
| $f_{L,tight}$ | $3.30 \times 10^{-1}$ | dimensionless | Fraction of lymph flow to tight tissues | [83] |
| $\sigma_{tight}$ | $9.45 \times 10^{-1}$ | dimensionless | Vascular reflection coefficient for tight tissues | [83] |
| $\sigma_{leaky}$ | $6.87 \times 10^{-1}$ | dimensionless | Vascular reflection coefficient for leaky tissues | [83] |
| $\sigma_L$ | $2.00 \times 10^{-1}$ | dimensionless | Vascular reflection coefficient for lymph | [83] |
| *Denosumab-specific parameters* |  |  |  |  |
| $MW_{deno}$ | $1.49 \times 10^2$ | kDa | Molecular weight of denosumab | [88] |
| $V_{p,deno}$ | $3.39 \times 10^{-2}$ | L | Apparent plasma volume for denosumab | estimated |
| $F_{deno}$ | $6.10 \times 10^{-1}$ | dimensionless | Denosumab bioavailability following SC administration | [85] |
| $k_{a,deno}$ | $1.60 \times 10^{-1}$ | 1/day | Absorption rate constant | estimated |
| $k_{e,deno}$ | $3.10 \times 10^{-2}$ | 1/day | Non-specific elimination rate constant for denosumab | estimated |
| $k_{off,deno}$ | $5.37 \times 10^{-1}$ | 1/day | Dissociation rate constant for denosumab–RANKL complex | [129] |
| $K_{D,deno}$ | $2.64 \times 10^1$ | pM | Equilibrium dissociation constant for denosumab–RANKL binding | [129,130] |
| $C_{RANKL}$ | $1.00 \times 10^2$ | pM | RANKL capacity | estimated |
| $\lambda_{RANKL}$ | 2.20 | 1/day | RANKL degradation rate constant | estimated |
| *Romosozumab-specific parameters* |  |  |  |  |
| $MW_{romo}$ | $1.49 \times 10^2$ | kDa | Molecular weight of romosozumab | [86] |
| $V_{p,romo}$ | $7.00 \times 10^{-2}$ | L | Apparent plasma volume for romosozumab | estimated |
| $F_{romo}$ | $8.10 \times 10^{-1}$ | dimensionless | Romosozumab bioavailability following SC administration | [86] |
| $k_{a,romo}$ | $5.31 \times 10^{-1}$ | 1/day | Absorption rate constant for romosozumab | estimated |
| $k_{e,romo}$ | $1.95 \times 10^{-1}$ | 1/day | Non-specific elimination rate constant for romosozumab | estimated |
| $k_{off,romo}$ | $2.51 \times 10^1$ | 1/day | Dissociation rate constant for romosozumab–SOST complex | [131] |
| $K_{D,romo}$ | 3.00 | pM | Equilibrium dissociation constant for romosozumab–SOST binding | [131] |
| $C_{SOST}$ | $2.13 \times 10^2$ | pM | Sclerostin capacity | estimated |
| $k_{int,romo}$ | $5.59 \times 10^{-2}$ | 1/day | Internalization rate of romosozumab–SOST complex | estimated |

$k_{on}$ values were calculated from the corresponding equilibrium dissociation constants and unbinding rate constants as $k_{on} = k_{off}/K_D$ . Internalization rate constant for denosumab–RANKL complex was assumed to be equal to the RANKL degradation rate constant. *PBPK* - *physiology-based pharmacokinetic*; *RANKL* - *receptor activator of nuclear factor κB ligand*; *SOST* - *sclerostin*.

**Table 4.**
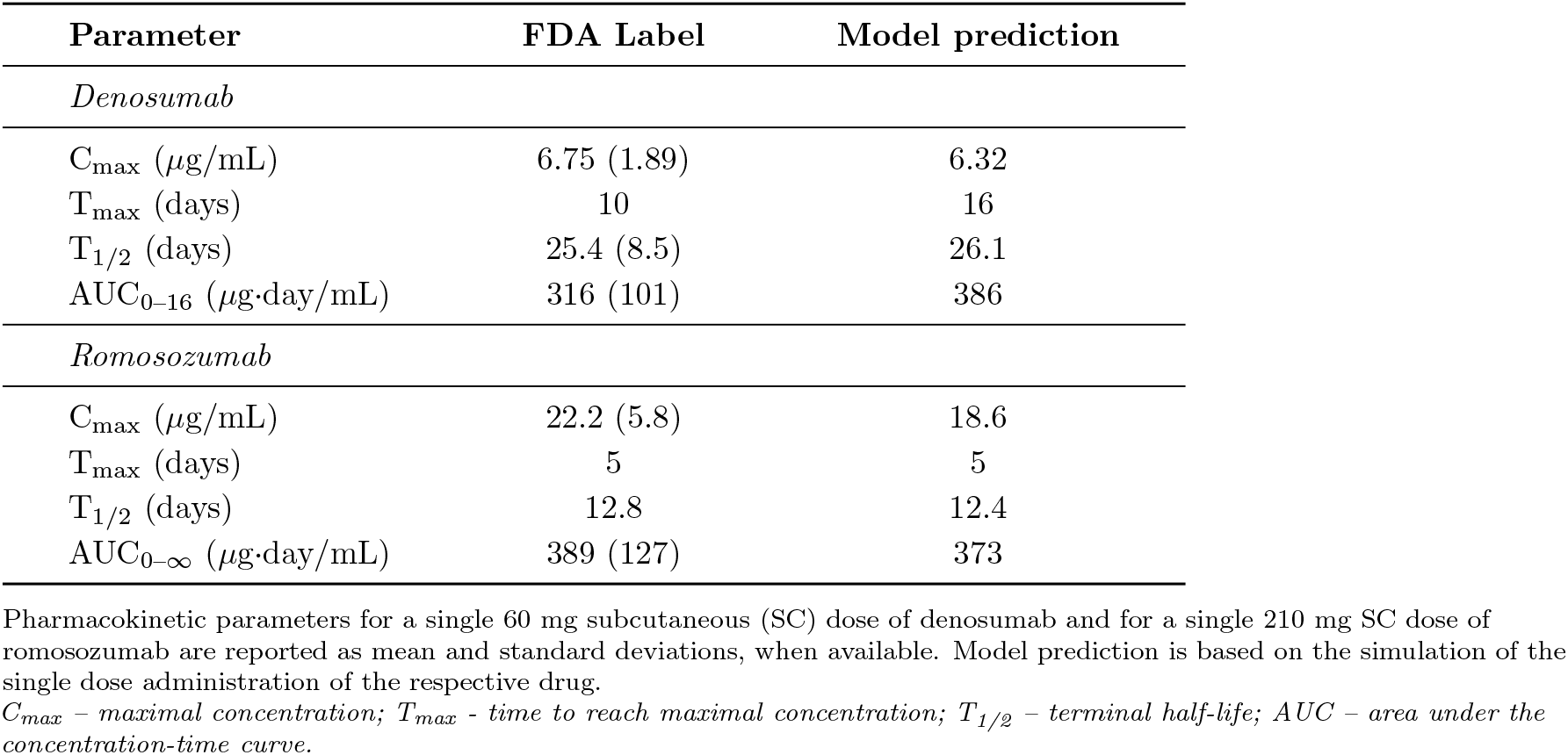
Pharmacokinetic parameters for denosumab and romosozumab.

### Model calibration and validation

The integrated bone remodeling model was calibrated to clinical data describing healthy postmenopausal bone loss and bone gain observed after treatment with denosumab, romosozumab, and micronized estradiol. Model performance was evaluated by its ability to reproduce longitudinal changes in BMD and BTMs across multiple interventions (Fig 4). Seven model parameters determining bone dynamics were selected for calibration and estimated by fitting the model sequentially to each intervention-specific dataset. In addition, the four parameters relating BMD dynamics to BTMs were fit to match the bone resoprtion or formation rate to the respective metabolite at baseline. Estimated parameter values are summarized in Table 5.

**Table 5.** Calibrated model parameters.

| Parameter | Description | Value | Unit |
| --- | --- | --- | --- |
| $\mathcal{D}_{OB_p}$ | Maximal rate of osteoblast progenitor maturation | $1.09 \times 10^{-2}$ | 1/day |
| $k_{form}$ | Bone formation rate constant | 3.01 | %/pM/day |
| $EC50_{Wnt}$ | EC50 value for Wnt signaling on osteoblasts | $1.22 \times 10^3$ | pM |
| $k_{on}^{SOST:LRP}$ | Binding rate constant for SOST–LRP5/6 complex formation | $4.40 \times 10^{-1}$ | 1/pM/day |
| $K_{AOC_a}^{TGF\beta}$ | Activation rate constant for TGF-β effect on osteoclast apoptosis | 2.70 | pM |
| $K_{AOC_a}^{Estr}$ | Activation rate constant for estrogen effect on osteoclast apoptosis | $5.26 \times 10^1$ | pM |
| $C_{OPG}$ | Initial OPG concentration | $2.00 \times 10^2$ | pM |
| $q^{CTX}$ | Coefficient relating bone resorption rate to CTX levels | $2.08 \times 10^1$ | dimensionless |
| $q^{NTX}$ | Coefficient relating bone resorption rate to NTX levels | $1.68 \times 10^3$ | dimensionless |
| $q^{BSAP}$ | Coefficient relating bone formation rate to BSAP levels | $1.20 \times 10^3$ | dimensionless |
| $q^{P1NP}$ | Coefficient relating bone formation rate to P1NP levels | $2.50 \times 10^3$ | dimensionless |

$EC50$ – half-maximal effective concentration; $SOST$ – sclerostin; $LRP$ – low-density lipoprotein receptor-related protein; $TGF\text{-}\beta$ – transforming growth factor $\beta$ ; $OPG$ – osteoprotegerin; $CTX$ – C-terminal telopeptide; $NTX$ – N-terminal telopeptide; $BSAP$ – bone-specific alkaline phosphatase; $P1NP$ – procollagen type I N-terminal propeptide.

**Fig 4.**
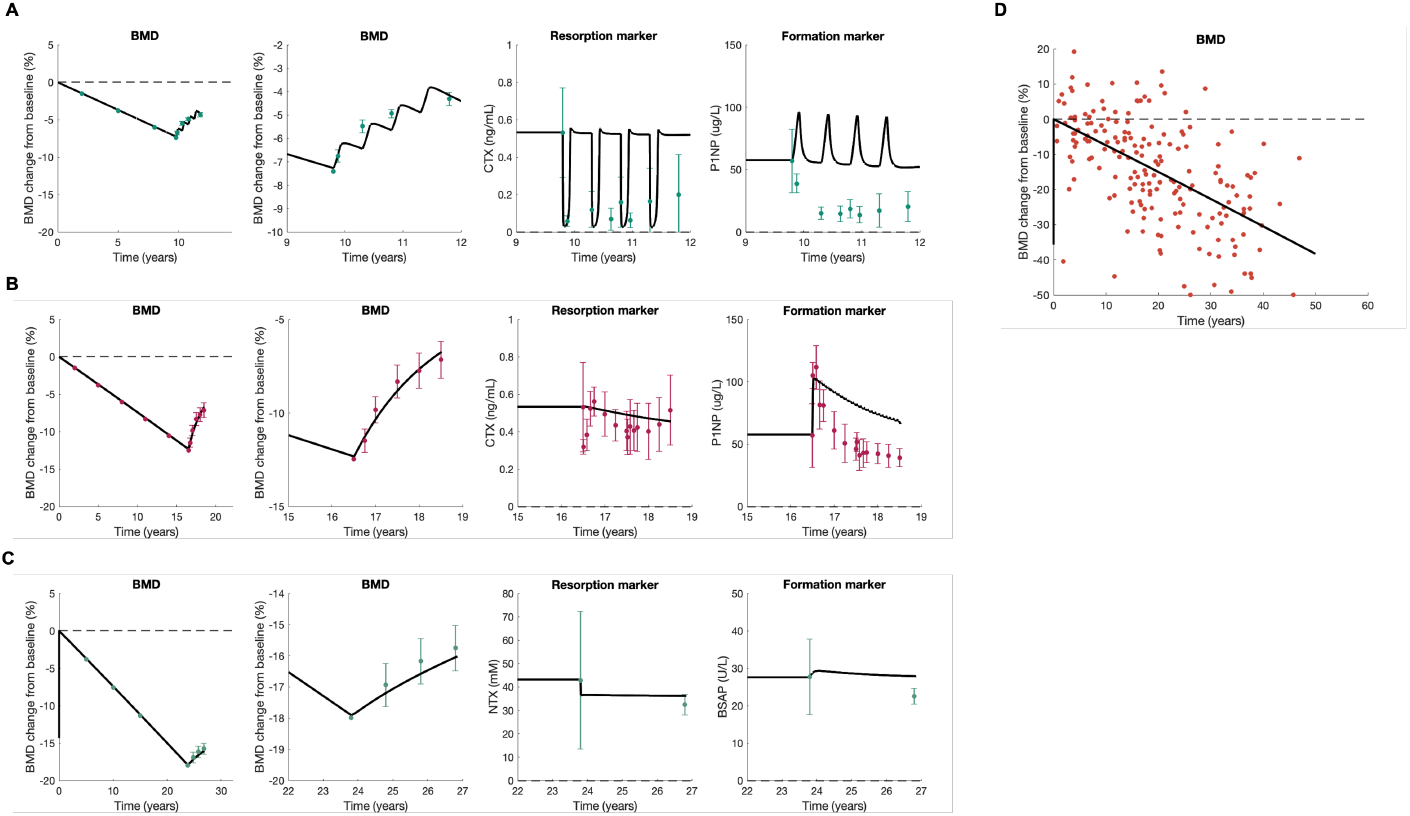
Bone remodeling model calibration. The bone remodeling model was calibrated to clinical data on bone mineral density (BMD) and bone turnover metabolite (BTM) dynamics after treatment with (A) 60 mg SC Q6M of RANKL inhibitor denosumab for 2 years; (B) 210 mg SC QM of sclerostin inhibitor romosozumab for 2 years; and (C) 0.25 mg PO QD of micronized estradiol for 3 years. Healthy postmenopausal decline in BMD was simulated for all 3 datasets from the average age of menopause onset to the average age at enrollment, followed by a simulation of the respective intervention. BMD, bone resorption marker and bone formation marker data for all three datasets were fit simultaneously. Experimental data reflect the study population mean and standard error reported in the respective clinical trial. (D) calibrated model simulation overlaid on individual-level BMD data in postmenopausal women.

Upon administration of bone-modifying agents, the model accurately reproduced the magnitude of BMD gain reported in clinical trials (Fig 4A–C). Treatment with 60 mg of denosumab SC Q6M for 2 years resulted in a cumulative BMD gain of 3.1%. Each dose of denosumab induced a rapid initial bone gain, followed by a stabilization phase and a rebound of bone resorption, reflecting potent suppression of osteoclast activity and of osteoclast pool after the drug is eliminated (Fig 4A). Administration of 210 mg of romosozumab SC QM for 2 years induced a pronounced initial increase in BMD, followed by a slower accrual phase, resulting in net BMD gain of 5.6% over the course of treatment (Fig 4B). Treatment with 0.25 mg of micronized estradiol QD led to modest recovery of BMD, with 1.9% gain over 3 years of therapy, reflecting partial suppression of bone resorption (Fig 4C). Following calibration, the base model was able to capture the steady decline in BMD observed in healthy postmenopausal women over a 50-year period following menopause onset (Fig 4D). The simulated rate of bone loss was 0.62% per year.

The model reproduced the qualitative dynamics of bone resorption markers across all interventions, including rapid 96% suppression of CTX following denosumab administration, steady decrease in CTX to 85% of the baseline level over the course of romosozumab treatment, and sustained 15% reduction in NTX after 3 years of estrogen supplementation. In contrast, the model was not able to fully replicate clinically-observed dynamics of bone formation markers. For romosozumab, the model successfully captured the initial increase in P1NP, followed by a gradual decline over time, although the magnitude of the decrease was underestimated relative to clinical data (Fig 4B). For denosumab, the model reproduced a steady decrease in P1NP during the course of treatment; however, the simulated decline was substantially smaller than clinically-observed values (Fig 4A). These discrepancies are likely due to a simplified representation of BTM dynamics, and the increased focus on BMD trajectories during calibration.

Overall, the calibrated model accurately captures long-term bone loss in healthy postmenopausal women and is able to reproduce the effect of pharmacological interventions. These results support the structural validity of the model, enabling assessment of the impact of genetic perturbations.

### Mechanism of estrogen regulation

To better understand the impact of different mechanisms of estrogen regulation on bone dynamics, we performed an ablation experiment in which estrogen effects were removed from the model one at a time. BMD trajectories and annual BMD loss rates were evaluated under normal postmenopausal estrogen levels and under a 90% estrogen suppression scenario (Fig 5).

**Fig 5.**
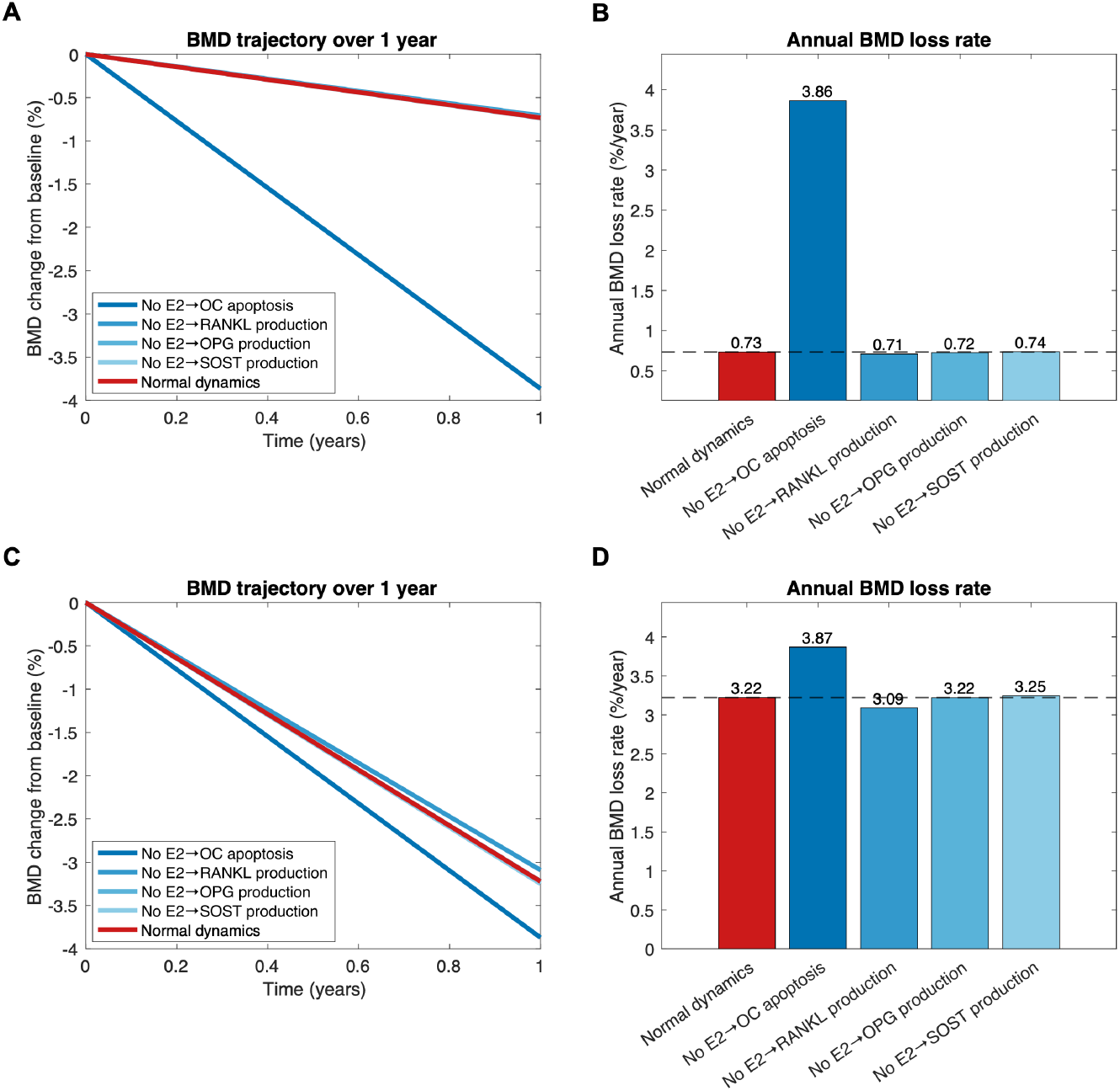
Estrogen effect ablation. Estrogen regulation of osteoclast apoptosis, RANKL production, OPG production, and SOST production were removed one at a time, and the bone mineral density (BMD) dynamics under ablation scenarios was compared to the BMD dynamics under the base model with all 4 effects included. Panels A and B display BMD dynamics and BMD loss rate for under normal postmenopausal estrogen levels. Panel C and D demonstrate BMD dynamics and BMD loss rate under further 90% suppression of estrogen. *E2 – estradiol; OC – osteoclast; RANKL – receptor activator of nuclear factor κB ligand; OPG – osteoprotegerin; SOST – sclerostin*.

Under normal postmenopausal estrogen conditions, ablation of estrogen’s effect on osteoclast apoptosis resulted in a 6-fold increase in the rate of BMD loss compared to the baseline scenario in which all regulatory mechanisms were intact (Fig 5A,B). In contrast, removal of estrogen regulation of RANKL, OPG, or sclerostin production did not affect BMD trajectories. Further suppression of estrogen resulted in a 4.5-fold increase in BMD loss rate when all regulatory mechanisms were included in the model (3.22 %/year vs 0.73 %/year in healthy postmenopausal state). Under the estrogen suppression scenario, removal of estrogen’s effect on osteoclast apoptosis remained the most influential perturbation, resulting in bone loss rate comparable to total estrogen depletion (3.87 % bone loss per year, Fig 5C,D). At the same time, removal of estrogen-mediated regulation of RANKL production had the opposite effect on BMD loss rate (3.09 %/year), while ablation of the effect on sclerostin production resulted in a slight increase in bone loss rate compared to the full model (3.25 %/year). Removal of OPG regulation by estrogen did not affect BMD dynamics. Together, these results suggest that estrogen primarily protects the bone by promoting osteoclast apoptosis, and prolonged osteoclast survival is the primary driver of bone loss under low estrogen conditions.

### Genetic effect estimation

For each SNP, we independently estimated genetic effect parameters by fitting the model to expected deviations in BMD dynamics from the reference trajectory estimated for each genotype. For variants in *TGFB1* and *CYP19A1*, we calibrated the model to the observed TGF-*β* and estradiol levels, respectively. The majority of the analyzed SNPs (19/22) occurred in regulatory regions of the gene, and thus were assumed to affect expression level of the corresponding protein. All 3 SNPs occurring in coding regions were missense mutations. *OPG* rs2073618 results in amino acid substitution from lysine to asparagine at position 3, which is located within the signal peptide sequence and is likely to affect OPG transport rather than its binding affinity [132–134]. Similarly, *TGFB1* rs1800470 is a missense variant resulting in lysine to proline substitution at codon 10 within the signal peptide, affecting TGF-*β* trafficking [135, 136]. Therefore, these variants were assumed to affect expression level of the corresponding proteins rather than binding. The missense variant in *LRP5* rs3736228 causes alanine to valine substitution at amino acid position 1330, which is located within the extracellular domain of the receptor and was assumed to affect LRP5 binding.

Across all variants, the calibrated model accurately replicated previously reported per-SNP effects (Fig 6). For SNPs affecting expression of *ESR1, CYP19A1, RANK, RANKL, SOST* and *LRP5*, the estimated effect sizes ranged from -0.07 to 0.10, which converts to 0.95–1.07-fold change in expression levels of the respective proteins Table 6. The variants in *OPG* were estimated to have stronger effects (*β*: -0.41–0.32), resulting in 0.75–1.25-fold differences in OPG expression. Similarly, the *TGFB1* variants were estimated to produce a larger effect on TGF-*β* expression, with each alternate allele of *TGFB1* rs1800469 decreasing TGF-*β* expression 1.6-fold. For the two SNPs in *LRP5*, the estimated effect was stronger for the variant affecting protein binding than for the variant affecting LRP5 expression (-0.27 vs -0.02). A summary of effect size estimates and their their 95% confidence intervals is provided in Table 6. Overall, our findings suggest that individual genetic variants produce modest effects on system parameters in either direction.

**Table 6.** Mechanistic effect estimates for included SNPs.

| Gene | SNP | Mutation | Mechanistic effect | Marginal $\beta$ | Adjusted $\beta$ |
| --- | --- | --- | --- | --- | --- |
| ESR1 | rs2234693 | T>C | Estrogen signaling sensitivity | -0.0401 [-0.00496 - -0.0306] | -0.0629 |
|  | rs9340799 | A>G | Estrogen signaling sensitivity | -0.0123 [-0.0184 - -0.0062] | 0.0288 |
|  | rs2228480 | G>A | Estrogen signaling sensitivity | -0.0466 [-0.0865 - -0.0067] | -0.0468 |
|  | rs9479055 | C>A | Estrogen signaling sensitivity | -0.0300 [-0.0400 - -0.0200] | -0.0300 |
|  | rs1999805 | G>A | Estrogen signaling sensitivity | -0.0232 [-0.0298 - 0.0166] | -0.0219 |
| CYP19A1 | rs727479 | C>A | Estrogen production | 0.1003 [0.0753 - 0.1253] | 0.0860 |
|  | rs4646 | A>C | Estrogen production | 0.0515 [0.0408 - 0.0622] | -0.0274 |
|  | rs10046 | G>A | Estrogen production | 0.0877 [0.0576 - 0.1178] | 0.0647 |
| RANK | rs884205 | A>C | RANK expression | -0.0150 [-0.0225 - -0.0075] | -0.0006 |
|  | rs3018362 | A>G | RANK expression | -0.0300 [-0.0413 - -0.0188] | -0.0297 |
| RANKL | rs9525638 | T>C | RANKL expression | -0.0352 [-0.0528 - -0.0176] | -0.0332 |
|  | rs9594738 | C>T | RANKL expression | 0.0316 [0.0176 - 0.0456] | 0.0293 |
| OPG | rs7839059 | C>A | OPG expression | -0.4120 [-0.5036 - -0.3204] | -0.7113 |
|  | rs4355801 | A>G | OPG expression | 0.3224 [0.1842 - 0.4606] | 1.9995 |
|  | rs3102735 | T>C | OPG expression | -0.0950 [-0.1900 - 0] | -0.2847 |
|  | rs2073618 | G>C | OPG expression | 0.1392 [0.0464-0.2320] | 1.5686 |
| SOST | rs7209826 | A>G | Sclerosin secretion | -0.0334 [-0.0451 - -0.0217] | -0.0244 |
|  | rs188810925 | G>A | Sclerosin secretion | -0.0658 [-0.0811 - -0.0505] | -0.0623 |
| LRP5 | rs3736228 | C>T | LRP5 binding | -0.2732 [-0.4024 - -0.0930] | -0.4437 |
|  | rs11228240 | C>T | LRP5 expression | -0.0234 [-0.0478 - 0] | 0.2636 |
| TGFB1 | rs1800470 | G>A | TGF-β trafficking | -0.2194 [-0.315 - -0.1422] | 1.0159 |
| | rs1800469 | A>G | TGF-β trafficking | -0.6384 [-0.9466 - -0.3892] | -1.4841 |

*SNP* - single nucleotide polymorphism; $\beta$ - genetic modifier parameter; *ESR1* - estrogen receptor 1; *CYP19A1* - aromatase; *RANK* - receptor activator of nuclear factor κB; *RANKL* - receptor activator of nuclear factor κB ligand; *OPG* - osteoprotegerin; *SOST* - sclerostin; *LRP5* - low-density lipoprotein receptor-related protein 5; *TGFB1* - transforming growth factor-β 1. Marginal effect sizes are presented as mean estimates and their 95% confidence intervals. Adjusted effect sizes represent linkage disequilibrium-adjusted values for the mean marginal estimate.

**Fig 6.**
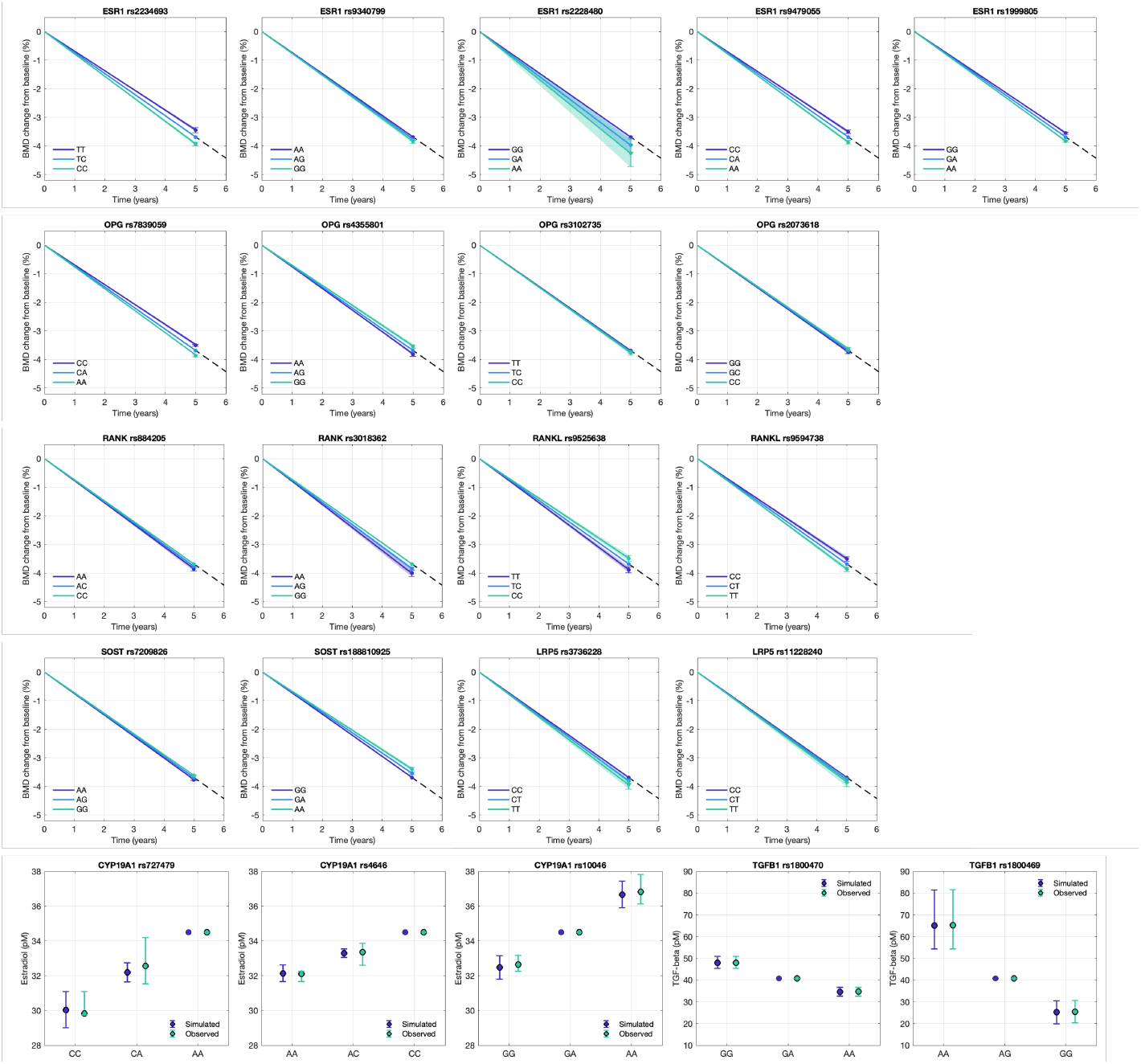
Genetic effect calibration. Parameters describing genetic effect were independently estimated for each single nucleotide polymirphism (SNP) to fit the observed effect on BMD (for SNPs in *ESR1, OPG, RANK, RANKL, SOST*, and *LRP5*) or on molecular concentrations (*CYP19A1*, and *TGFB1*). The base model simulation was assumed to occur for the The expected genotype based on global allele frequencies was assumed to follow the bone mineral density (BMD) trajectory simulated by the base model, and genotype-specific deviations from the base trajectory were calculated based on effect size reported in previous genetic association studies. Error bars represent 95% confidence intervals in reported effect sizes. Shaded areas indicate 95% prediction intervals. *ESR1 – estrogen receptor α; OPG – osteoprotegerin; RANK – receptor activator of nuclear factor κB; RANKL – receptor activator of nuclear factor κB ligand; SOST – sclerostin; LRP5 – low-density lipoprotein receptor-related protein 5; CYP19A1 – aromatase; TGFB1 – transforming growth factor-β 1*.

The estimated marginal effect sizes were adjusted for LD using pairwise correlation between the SNPs obtained from 1000 Genomes Project. In most loci, LD adjustment resulted in slight attenuation of marginal effect sizes with no change in the direction of the effect. Due to high correlation between the included *RANK* polymorphisms, rs3018362 almost entirely captured the shared genetic signal, resulting in LD-adjusted effect estimate for rs884205 that was close to zero. The direction of effect was reversed after LD adjustment for *ESR1* rs9340799 and *CYP19A1* rs4646, while their magnitudes remained similar to the unadjusted estimates. Due to high correlation, the LD-adjusted effects in *OPG* SNPs were largely inflated compared to the marginal estimates, with rs2073618 and rs4355801 producing 3- and 4-fold increase in expression levels after the adjustment, respectively (Table 6). A similar trend was observed for *TGFB1* variants, where LD-adjustment resulted in 2-fold increase in the effect size for rs1800469, and 5-fold increase in rs1800470 effect, while also reversing the effect direction for the latter.

### Population modeling

To evaluate the combined effect of multiple genetic polymorphisms on bone dynamics, we simulated BMD trajectories in a virtual population of 1,000 individuals that differed by ancestry and genetic covariates. When fixed genetic effects were used to simulate BMD trajectories in the population, the median 5-year bone loss was predicted to be 3.56% [IQR: 2.87–4.18%, 95% interval: 1.54–5.19%]. The simulated distribution captured approximately 22% of the assumed variance in BMD loss. Incorporation of uncertainty in genetic effect estimates slightly expanded prediction interval, resulting in the median 5-year BMD loss of 3.57% [IQR: 2.90–4.22%, 95% interval: 1.31–5.24%]. The simulated population distribution captured a range of literature-reported values, and accounted for 25% of the assumed variance in the outcome (Fig. 7A).

**Fig 7.**
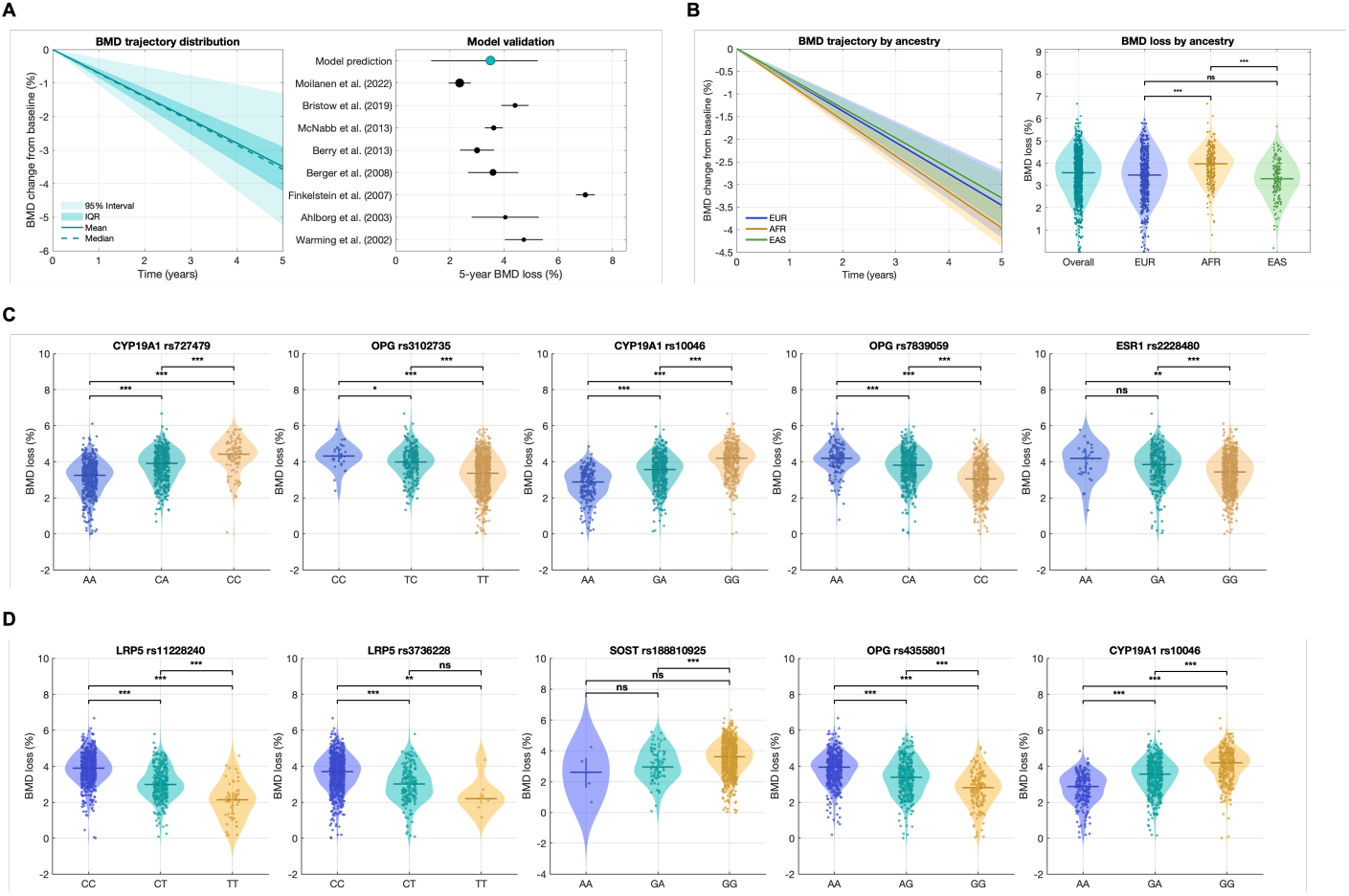
Population modeling. The combined effect of genetics was modeled by simulating bone mineral density (BMD) dynamics in a virtual population (N = 1,000) for 5 years. Panel A displays the distribution of simulated BMD trajectories and compares the predicted 5-year BMD loss to the values for postmenopausal bone loss reported in the literature. Panel B shows the distribution (mean and 95% confidence intervals) of BMD trajectories stratified by genetic ancestry. Panels C and D present top 5 single nucleotide polymorphisms (SNPs) with risk-enhancing or protective effect, respectively, as defined by the median bone loss over 5 years in the individuals with the homozygous genotype. *IQR – interquartile range; EUR – European ancestry; AFR – African ancestry; EAS – East Asian ancestry; CYP19A1 – aromatase; OPG – osteoprotegerin; ESR1 – estrogen receptor α; LRP5 – low-density lipoprotein receptor-related protein 5; SOST – sclerostin *p-value < 0*.*05; **p-value < 0*.*01; ***p-value < 0*.*001; ns – non-significant*

To better understand how genetic differences across populations affect bone dynamics, we stratified our analysis by genetic ancestry. Individuals of African ancestry were predicted to experience significantly higher BMD loss over 5 years following the menopause onset compared to their European and East Asian counterparts (AFR: 3.86%, EUR: 3.46%, EAS: 3.28%, *p <* 0.001, Fig. 7B).

To evaluate the effect of individual SNPs on BMD dynamics in the virtual population, we examined genotype-specific distributions of BMD loss for each SNPs (Fig. 7C,D). SNPs in *CYP19A1* and *OPG* were associated with highest BMD loss, while variants in *LRP5* produced the strongest protective effect in the virtual population. Virtual subjects homozygous for alternate allele (CC) in *CYP19A1* rs727479 were predicted to experience the highest bone loss over 5 years (4.42%, 95% interval: 2.18–5.78%), followed by *OPG* rs3102735 CC (4.32%, 95% interval: 2.82–5.40%) and *CYP19A1* rs10046 GG (4.20% [95% interval: 2.13-5.55%]. Homozygous alternate genotypes in both *LRP5* SNPs, rs11228240 (TT) and rs3736228 (TT), were associated with the lowest bone loss over 5 years (2.14% [95% interval:0.18-4.01%] and 2.20% [IQR: 1.25-4.18%], respectively). Across all variants, we observed a weak effect of individual SNPs on BMD dynamics and a wide distribution of BMD loss within each genotype group, suggesting that interactions between SNPs play a crucial role in extreme phenotypes.

### Genetic interaction analysis

To identify the epistatic effects between the SNPs included in the model, for each pair of SNPs, we fit a regression model including an interaction term. Most SNPs had little to no interactions with other variants. However, when the interaction was observed, most of the interactions were such that the combined effect of the SNPs was attenuated. The strongest interaction effects occurred between the SNPs located within the same locus and in LD with each other (*OPG* r4355801, rs3102735, rs2073618, and *SOST* rs7209826 and rs188810925, Fig. 8A). *SOST* rs188810925, *OPG* rs3102735, *CYP19A1* rs727479 and *TGFB1* rs1800469 were among the variants with the most cross-gene interactions. *ESR1* rs9479055 and *SOST* rs7209826 had the largest synergistic effect for the variants not located in the same gene (0.28, standard error (SE): 0.06), followed by the interactions between *CYP19A1* rs727479 and *SOST* rs7209826 (0.26 SE: 0.06), and *ESR1* rs9479055 with *SOST* rs7209826 (0.23, SE: 0.06). The mean BMD loss for pairwise combinations of *ESR1* rs9479055, *SOST* rs7209826 and *CYP19A1* rs727479 genotypes are presented in Figure 8B. *ESR1* and *SOST* SNPs interact to modify each other’s effect, such that the relationship between *ESR1* rs9479055 genotype and BMD loss depends on *SOST* rs7209826 genotype, and vice versa. For the interaction between *CYP19A1* rs727479 and *SOST* rs7209826, the direction of *CYP19A1* effect remained consistent across *SOST* genotypes; however, its effect was attenuated in virtual individuals carrying GG genotype at *SOST* rs7209826. The strongest cross-gene interactions producing sub-additive combined effect was detected between *RANK* rs3018362 and *SOST* rs188810925 (-0.29, SE: 0.17), and between *SOST* rs188810925 and *CYP19A1* rs4646 (-0.26, SE: 0.18), although it is important to note that only a small fraction of individuals in our virtual cohort caried AA genotype at *SOST* rs188810925 (n = 4). Bone-protective effect of RANK was only detectable in individuals carrying a risk allele at *SOST* rs188810925 (Fig. 8C). At the same time, *SOST* rs188810925 was not able to fully rescue the bone-destructive effects of *CYP19A1* rs4646. These findings demonstrate that while most genetic effects are additive, non-linear interactions are common, highlighting the importance of mechanistic modeling in determining the combined genetic burden.

**Fig 8.**
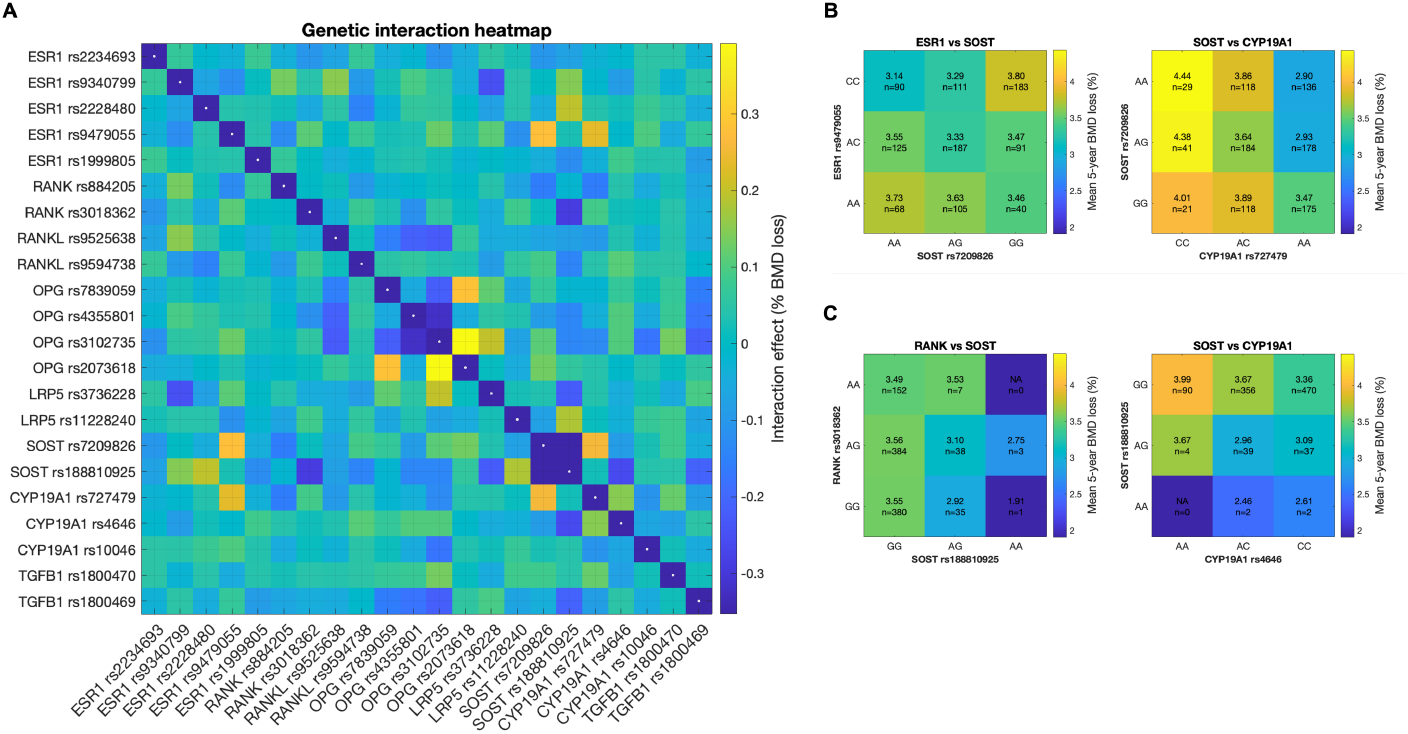
Genetic interaction analysis. The epistatic effect between a pair of SNPs was estimated by fitting an additive genetic model (panel A). Positive interactions indicate synergistic effects between SNPs, while negative interactions suggest attenuation of additive effects. The magnitude of the effect reflects the deviation from additivity in units of percent BMD loss. Panel B displays the mean BMD loss in each genotype combination for the SNPs located within different loci with highest positive interaction effect. Likewise, panel C displays the mean BMD loss for the SNPs with the highest negative interaction effect. *SNP – single nucleotide polymorphism; BMD – bone mineral density; ESR1 – estrogen receptor 1; CYP19A1 – aromatase; RANK – receptor activator of nuclear factor κB; RANKL – receptor activator of nuclear factor κB ligand; OPG – osteoprotegerin; SOST – sclerostin; LRP5 – low-density lipoprotein receptor-related protein 5; TGFB1 – transforming growth factor-β 1*.

## Discussion

In this study, we introduce a novel mechanistic modeling framework to evaluate the impact of genetic variability on bone remodeling in postmenopausal women. While several mathematical models of bone remodeling exist in the literature, only a few of the existing frameworks explicitly model regulatory effects of estrogen [20–25, 137]. Furthermore, previous models differed in the assumed mechanisms that drive estrogen effect. In the present study, we integrated several mechanisms of estrogenic regulation of bone remodeling described in previous studies and evaluated the relative contribution of each mechanism. We further expand on the work of others by mechanistically incorporating genetic variants known to affect bone remodeling and investigating the complexity of their combined effect. As our model supports incorporating patient-specific genetic covariates and simulating personalized BMD trajectories, we hope that our proposed framework will help bridge the gap between scientific discovery and clinical practice, and enable personalized prediction of bone loss risk.

Our model was initially parameterized using the literature-informed parameter values. However, we observed a large variability in reported estimates across studies. To further constrain the parameter space, we introduced relationships between the parameters, which were more consistent in the literature. We further calibrated the model using observational cohort study data on healthy BMD decline in postmenopausal women, and clinical trial data on BMD gain observed after treatments with RANKL inhibitor (denosumab), sclerostin inhibitor (romosozumab) and micronized estradiol. Since denosumab targets the catabolic pathway, romosozumab targets the anabolic pathway, and micronized estradiol modulates hormonal regulation, these three interventions allowed us to effectively estimate a unique set of parameters describing the corresponding part of the system. Despite well-recognized limitations of BTMs, such as substantial inter-individual variability and weak predictive power, these markers provided additional information about therapeutic effects on bone formation and resorption rates [69, 138–140]. These insights were used to further inform parameter selection for calibration. The use of drug response outcome data for model calibration required accurate mechanistic representation of interventions themselves. Thus, we built mPBPK models with full TMDD for denosumab and romosozumab, which allowed us to expand the set of calibrated parameters and estimate target capacities for RANKL and sclerostin, as well as RANKL internalization rate constant. Our mPBPK models were able to accurately capture the data across a range of doses and produced the PK metrics that closely matched the ones reported in FDA labels for the clinically administered doses.

The calibrated bone remodeling model accurately described the BMD dynamics following denosumab and romosozumab treatments for 2 years and estrogen supplementation for 3 years. While romosozumab and estrogen supplementation provided a steady increase in BMD over time with a reduction in BMD gain rate after several years of therapy, each dose of denosumab produced a rapid BMD gain followed by stabilization and a period of enhanced resorption prior to subsequent dose. The model captured NTX and CTX dynamics relatively well across the interventions, slightly underestimating suppression of bone resorption in individuals treated with micronized estradiol. On the other hand, the model produced poor fits to P1NP and BSAP data, although we were able to capture the general direction of effect on the biomarker. Due to high variability in BTM levels, our calibration setup prioritized BMD data over BTM dynamics, which may have affected the final fits. In addition, the inability of the model to quantitatively describe changes in bone resorption markers may be in part explained by our simplified approach to model BTM dynamics. Choosing a different structural form to relate bone resorption and formation rates to the respective biomarkers, or mechanistically representing BTM production and degradation rates may result in improved fits, while increasing model complexity. Overall, the multilevel calibration strategy we used enabled our model to accurately capture BMD dynamics across various interventions and demonstrated the structural validity of the model, providing a strong foundation for incorporation of genetic variability.

For genetic effect integration, we selected SNPs in genes that could be mechanistically represented within the modeling framework. This limited the pool of candidate genes to the ones encoding the proteins represented as species in our model, or their receptors. While our selection included the genes in key regulatory pathways, such as RAN-RANKL-OPG, Wnt, TGF-*β* and estrogen signaling, we were not able to mechanistically incorporate well-replicated SNPs in genes in other pathways (e.g., *MPP7, RUNX2, SMAD, RSPO3*) [29, 31, 103]. More complex models incorporating additional mechanistic details and signaling pathways could be developed in the future to explore the role of genetic polymorphisms in these genes. Importantly, the objective of our model was to demonstrate the proof-of-concept on mechanistic integration of genetic effects, rather than to model the overall genetic variability. The combined effect of the 22 SNPs included in the model accounted for only 25% of the overall variability in BMD loss, suggesting that non-genetic factors (e.g., clinical, demographic, socio-behavioral) play a major role in determining bone dynamics in postmenopausal women. Incorporating additional genetic factors and non-genetic covariates should be an important focus on future research.

Mathematically, genetic effect was incorporated as a fold change in model parameters, such as production rate of the species, its binding activity, or signaling sensitivity, based on the location of the SNP within the gene. While quantitative fold changes in RNA or protein expression levels relative to baseline expression for each genotype would be desirable to enable a fully mechanistic representation of genetic effects from functional perspective, the expression levels are highly variable across tissues and across studies [141]. Furthermore, bone tissue is not included in the Genotype-Tissue Expression (GTEx) analysis, requiring additional assumptions for estimation of expression levels in the bone [141]. Therefore, in this study, we used GWAS data to estimate the parameters representing genetic modifiers on base model parameters. Importantly, GWAS-reported per-SNP effect size estimates represent association strength between the SNP and the outcome and do not imply causal effect. Therefore, we interpret the estimated genetic modifier as the fold change in the respective model parameter producing the BMD change observed in GWAS for a given SNP, rather than a fold change in expression level or affinity. This approach makes our model less mechanistic, but provides a direct link from the genetic variant to clinical outcome. As GWAS effect sizes are reported in standard deviation units, we made an assumption on the standard deviation of BMD loss in postmenopausal women. The assumed standard deviation of 2% BMD loss over 5 years spans the data used to compare the virtual population predictions to the real-world values. Importantly, the fraction of variance explained by genetics is independent of this assumption.

Upon calibration of marginal effect sizes for each SNP, our model was able to: (1) capture healthy BMD decline in postmenopausal women; (2) accurately reproduce BMD gain following pharmacological interventions affecting different parts of the system; and (3) replicate GWAS-reported shift in BMD for each SNP independently. Successful replication of clinical outcome data for these individual perturbations increased confidence in the structural validity of the model and supported its extension to scenarios involving combinations of genetic perturbations. To evaluate the combined effect of multiple polymorphisms present at the same time, we generated a virtual population of 1,000 patients differing by ancestry and genetic covariates. To accurately represent a real-world population, we sampled genotypes, such that they matched ancestry-specific allele frequencies and LD-patterns among the included SNPs. Expectedly, the distribution of BMD trajectories simulated in the virtual population centered at the base model trajectory. The majority of independent observational studies we reviewed (7/8) reported the 5-year BMD decline values within the 90% prediction interval by our model (Figure 7A).

Overall, individual SNPs were weakly correlated with 5-year BMD loss, and there was a large variability in the simulated outcome among the virtual individuals with the same genotype at a given locus. This finding is in agreement with the general understanding of postmenopausal bone loss as a polygenic trait, further emphasizing the importance of combining multiple SNPs into models. Furthermore, the analysis of epistatic interactions suggested that many SNPs interact non-linearly, either enhancing or canceling each other’s effects. In some cases, the presence of a specific genotype at one locus was required for the other SNP to produce its effect (e.g. *RANK* rs3018362 and *SOST* rs188810925, Figure 8C), or the direction of effect for one SNP was dependent on the genotype at another locus (e.g., *ESR1* rs9470955 and *SOST* rs7209826, Figure 8B). Further work is needed to better characterize the molecular and cellular dynamics producing such interactions. Improved understanding of cross-gene and cross-pathway interactions may help identify novel therapeutic targets and inform new intervention strategies in the future.

Our simulations suggested disparities in bone loss across ancestries, with individuals of African ancestry experiencing greater average BMD loss over the same period than their European and East Asian counterparts. Literature provides limited data on BMD dynamics across ancestries. In general, osteoporosis is more prevalent among women ethnically identifying as Chinese or Japanese, and lower in women self-identifying as African American. However, these disparities have been suggested to mainly originate from differences in peak BMD rather than BMD decline, where the rates of bone loss are comparable across ethnicities. Furthermore, there is a poor correlation between self-reported ethnicity and genetic ancestry, and additional socio-economic and behavioral factors play an important role in determining bone dynamics, complicating the comparison of our ancestry-based predictions with reports stratifying outcomes by race or ethnicity. Our study suggests that based on genetics alone, individuals of African ancestry may be more prone to accelerated bone loss after the menopause. It would be interesting to explore the socio-behavioral factors that may play a role in mitigating these effects in future research.

Our study has several limitations. First, our cell population dynamics model does not account for the microstructural differences between cortical and trabecular bone and between bone at different sites. For model calibration and validation, we used clinical data reported for total hip BMD. In a similar way, our framework enables modeling BMD dynamics at other sites such as lumbar spine or femoral shaft, which can be a focus of future analyses. Second, our model does not integrate mechanical feedback or immune regulation of bone remodeling. Incorporation of mechanical stimulus has been widely implemented in recent bone remodeling models and is important for accurate representation of sclerostin dynamics, while immune cells and cytokines have been shown to regulate osteoclast differentiation. Since the focus of this study was on the genetic regulation of bone remodeling, which is unaffected by mechanical stimulus and the immune system, we selected a simpler model structure. However, inclusion of these components would enable analysis of disease-disease interactions (e.g., osteoporosis with rheumatoid arthritis or diabetes) and additional interventions (e.g., exercise), and should be a focus of future research. Third, despite our efforts to reduce model complexity, the final model structure is high-dimensional and includes 64 parameters. Only a limited number of parameters are identifiable given the available clinical data; thus, we relied on literature-reported parameter values, some of which had high variability in estimates across studies. Further efforts are needed within the field to summarize the available data, establish the standards for parameter extraction and quality control checks. Fourth, the genetic modifier parameters were estimated from GWAS-reported effect sizes rather than from functional genomics studies. To enable fully mechanistic integration of genetic effects, future studies should focus on incorporating transcriptomic and proteomic data for regulatory variants and variant effect prediction tools, such as Ensembl and Alphafold, to predict changes in protein affinity for non-synonymous missense mutations [116, 142]. In addition, biases present in the GWAS used to estimate the genetic effects, such as population stratification, population and publication bias, were propagated to the current study. To mitigate some of these limitations, we incorporated variants supported by multiple studies and accounted for population-specific LD structure when calculating genetic effects. In addition, only a portion of genetic variability was modeled in this study, and we did not model variability due to non-genetic factors. The objective of this work was to demonstrate a proof-of-concept for mechanistic integration of genetics within mathematical models of biological systems, rather than to comprehensively describe inter-individual differences in postmenopausal osteoporosis progression. Therefore, we did not incorporate additional variability. However, incorporating additional covariates would be an important next step to better describe the heterogeneity of bone dynamics in postmenopausal women and to enable personalized predictions of bone loss. The same framework for covariate incorporation that we used in this study can be generalized to non-genetic covariates. Lastly, model evaluation was limited to population-level data and did not include validation of genetic effects on independent datasets. To increase the confidence in model predictions, it would be important to investigate the model performance on individual-level data.

## Conclusion

In this study, we developed a novel mechanistic model of bone remodeling in postmenopausal women that explicitly incorporates estrogen signaling and genetic variability. The established framework is able to reproduce healthy BMD decline in postmenopausal women, clinical responses to catabolic, anabolic and hormonal therapies, and the effect of genetic perturbations on bone remodeling. Our model enables prediction of a combined effect of several genetic variants, and evaluation of non-linear genetic interactions from the mechanistic perspective. Although incorporation of additional covariates and individual-level validation will be necessary before clinical application, this work provides a proof-of-concept for integrating genetic risk factors into mechanistic models of disease. This approach can be extended to other conditions, and can be particularly useful for modeling diseases with high heritability and polygenic inheritance. We hope that our proposed framework will help bridge the gap between scientific discovery and clinical practice and enable personalized disease risk assessment.

## Data Availability

All data produced in the present study are available upon reasonable request to the authors

## AI Use Statement

During the preparation of this work, the author used ChatGPT to trouble shoot code in MATLAB used for virtual population sampling and data visualization. This tool has only been used in accordance with the Johns Hopkins University guidelines for responsible use of AI and appropriate references have been added. I have reviewed and edited the content as needed and I take full responsibility for the content of this manuscript.

